# PHIHDL: A Novel HDL Index Predicting Baseline Pulmonary Hemodynamics and Long-Term Survival in PAH

**DOI:** 10.64898/2026.08.31.26361587

**Authors:** Sebastian Pritz, Natalie Bordag, Vasile Foris, Valentina Biasin, Helga Billensteiner, Hansjörg Habisch, Tobias Madl, Gunther Marsche, Chandran Nagaraj, Susanne Suessner, Gabor Kovacs, Gustavo A. Heresi, Ulrich Bodenhofer, Horst Olschewski, Andrea Olschewski

## Abstract

**Rationale:** Pulmonary hypertension is defined by pulmonary hemodynamics, but diagnostic and prognostic biomarkers remain limited. Nuclear magnetic resonance (NMR) spectroscopy provides detailed insights, particularly in the lipid metabolism.

**Objectives:** To explore circulating NMR-derived metabolites and lipoprotein-related parameters for their association with pulmonary hemodynamics and to analyse their prognostic properties in pulmonary arterial hypertension (PAH).

**Methods:** Retrospective analysis of a PAH cohort with complete diagnostic workup including right heart catheterization and baseline serum samples, from the prospective GRaz Pulmonary Hypertension-Metabolism (GRAPH-M) registry.

**Measurements:** NMR-derived metabolites and lipoprotein-related parameters were analyzed for their association with clinically relevant parameters of PAH. We defined PHIHDL, a score derived from high-density lipoprotein (HDL) related measures based on their strong association with pulmonary hemodynamics, and evaluated its prognostic value.

**Results:** We included 100 patients with PAH treated at the PH clinic of LKH University Hospital, Medical University of Graz, between 2011 and 2021. Age was 61±15 years, female/male ratio 2.5, BMI 26±7 kg/m^2^, mPAP 41±16 mmHg, PAWP 8.8±3.2 mmHg, PVR 8.0±4.9 WU, and median survival was 8.0 years. During follow-up, 46 patients died. We identified a cluster of 12 HDL-related measures that showed significant inverse association to pulmonary hemodynamics and derived PHIHDL from the reversed scaled average of these particles. PHIHDL was associated with all-cause mortality after adjustment for age and sex (HR 2.96, 95% CI 1.52-5.70), independent of the clinical risk scores COMPERA 2.0 and REVEAL Lite 2.

**Conclusion:** PHIHDL, a pulmonary hemodynamics-based metabolomic score, provides independent prognostic information beyond established risk scores in PAH.

## Introduction

Pulmonary arterial hypertension (PAH) is associated with significantly increased morbidity and mortality, even when pulmonary pressure is only mildly elevated. (1) Important pathological mechanisms within the small pulmonary arteries have been identified, enabling the development of effective therapies targeting the prostacyclin, endothelin, and nitric oxide pathways (2), and several causal gene variants have been identified, (3). However, the systemic manifestations of PAH remain incompletely understood.

PAH is associated with abnormalities in circulating white blood cells (4–7), red blood cells (8), and metabolic pathways including elevated uric acid (9), and changes in lipid metabolism that resemble insulin resistance (10), although insulin sensitivity per se appears to be normal (11,12). In addition, elevated concentrations of specific fatty acids (13) and reduced levels of HDL cholesterol (14,15) have been linked to all-cause mortality in PAH patients.

NMR spectroscopy enables comprehensive profiling of circulating metabolites and lipoprotein-related parameters. In a UK cohort, reduced plasma levels of small HDL particles (HDL4) were directly associated with poor clinical outcomes in PAH patients (16), suggesting a potential link between HDL metabolism and disease pathobiology. We used NMR-based metabolomics and lipoprotein-related parameters to identify circulating metabolites associated with pulmonary hemodynamics in PAH. We identified a lipoprotein-related cluster comprising constituents of larger HDL particles (HDL1 and HDL2) that showed stronger associations with mortality than pulmonary hemodynamic measures.

## Methods

### Study design and cohorts

We prospectively enrolled patients into our Graz PH-Metabolism registry (GRAPH-M) at the Division of Pulmonology, Department of Internal Medicine, at the Medical University of Graz, who received right heart catheter diagnostics due to PH or a significant risk of PH. For this study, we included patients newly admitted to our PH clinic with Group 1 PAH, according to current guidelines (17), who underwent a complete diagnostic work-up for PH, including right heart catheterization (RHC), ventilation/perfusion scan, chest computed tomography, polysomnography, and screening for autoantibodies and HIV, between March 2011 and December 2021. RHC was clinically indicated, based on high pre-test probability for PH (significantly elevated systolic pulmonary arterial pressure (SPAP) in the Doppler echocardiography, or dilated pulmonary arteries, or elevated BNP, or poor 6 MWD, or cardiopulmonary exercise test, or unexplained dyspnoea) or a previous RHC. The diagnosis of PAH with a mean pulmonary arterial pressure >20 mmHg, PAWP≤15mmHg and PVR>2 WU, and exclusion of other forms of PH was confirmed by at least two experienced clinicians. All subjects provided written informed consent, and the study protocol was approved by the Ethics Committee of the Medical University of Graz (EK: 23-408 ex 10/11). Patients were excluded, if no baseline blood sample was available within 3 months of RHC or if there were no follow-up visits after RHC. Patients were also excluded if they had an incomplete diagnostic evaluation, no PH, or PH other than Group 1 PAH.

In most cases, blood was obtained from the superior vena cava after placement of the right heart catheter. In the other cases, blood was drawn from a peripheral vein. Serum was separated according to local standards and sample aliquots were stored at-80 °C at the Biobank Graz after written informed consent.

### NMR metabolomics

Nuclear magnetic resonance (NMR) spectra of sera were recorded on a Bruker Avance Neo 600 MHz spectrometer (Bruker GmbH, Ettlingen, Germany) as previously described. (18) Quantification of serum metabolites and lipoprotein subclasses from ^1^H-NMR spectra was performed using Bruker IVDr software, specifically B.I.Quant-PS 2.0.0 for the quantification of 41 serum metabolites in mmol/L units, and B.I.LISA (Lipoprotein Subclass Analysis) for the quantification of 112 serum lipoprotein parameters. Lipids are provided in units of mg/dL, apolipoproteins and particle numbers in nmol/L, ratios are dimensionless. All data is provided in Supplementary Data 1.

### Statistical Analysis

Continuous variables were assessed for normal distribution using visual inspection and the Shapiro-Wilk test. Normally distributed data are presented as mean ± standard deviation (SD), whereas skewed continuous variables are reported as median [Q1, Q3]. Categorical values are represented as ratios (A:B). Survival-related metrics, including hazard ratios (HR), are reported with 95% confidence intervals (CI).

Statistical analysis was performed using R version 4.3.3 (R Foundation for Statistical Computing, Vienna, Austria). All metabolic and lipoprotein-related parameters were subjected to a quality control preprocessing pipeline. They were subsequently filtered based on completeness (<20% missing values) to ensure sufficient sample size for robust correlation estimates. To reduce skewness and aid visualization, non-lipoprotein metabolic parameters were log_10_-transformed. Following transformation, principal component analysis (PCA) (19) was used to detect multivariate sample outliers identifying and excluding one sample (see Supplementary Note 1).

To account for multiple testing, p-values were adjusted using the Benjamini-Hochberg procedure. (20) To enrich for clustered associations, the correlation matrix was filtered bidirectionally by applying a scree-like heuristic (21), where the cutoff was set at the first major inflection point (the ‘knee’), to the count of metabolic and lipoprotein-related parameters remaining after filtering (y-axis) as a function of the minimum number of significant correlations required for each parameter, and vice versa (Fig. S1).

Pairwise Spearman rank correlations were calculated between the pre-processed NMR data and a pre-defined set of clinical parameters shown in Table 1 and Table S1 to capture the most robust associations in a framework of non-normal distributions and the discrete nature of lower-resolution parameters. We visually identified a prominent lipoprotein-related cluster, consisting of 12 HDL-related measures that were strongly associated with pulmonary hemodynamics. To facilitate the diagnostic evaluation of this cluster, we calculated a composite score, using these HDL-related measures (see Supplementary Note S2 and S3). We defined PHIHDL as the ‘Pulmonary Hypertension Index from High Density Lipoproteins’ and accounted for the inverse relationship to the hemodynamics by inverting the scale (1 – score). Because of the significant correlation of low HDL values with increased mortality, this resulted in higher values of PHIHDL being associated with increased mortality.

**Table 1:**
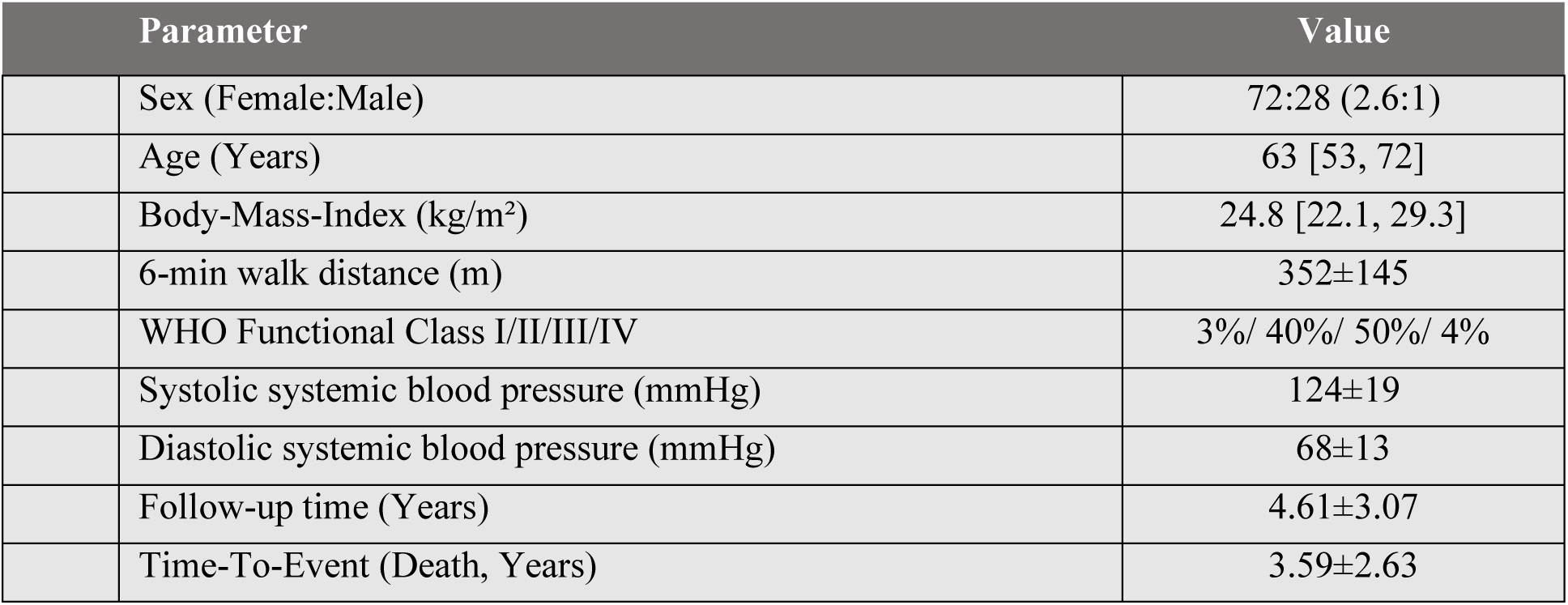
Patients’ physical characteristics (n=100) and observation times (mean±SD / median [quartiles])

We evaluated the prognostic utility of PHIHDL by (i) assessing its performance alone, ii) in combination with established tools (COMPERA2, REVEAL Lite 2) and (iii) testing the scaled replacement of NT-proBNP in these clinical scores.

For survival analysis, Kaplan-Meier curves were utilized. Cox proportional hazards models were used to determine hazard ratios for covariates of interest. To avoid data-driven cutpoint selection and minimize bias, numeric factors were dichotomized, i.e. categorized into low and high groups based on each factor’s baseline cohort median, and categorical versions were also dichotomized by aggregating groups (e.g., low and intermediate-low combined). In the hazards models, we adjusted for age and sex either one by one, or simultaneously. Models were developed for standalone scores, combined risk frameworks, and modified configurations incorporating PHIHDL. Whenever composite scores were compared to the original COMPERA2 or REVEAL Lite 2, to determine the incremental predictive value of the metabolic/lipoprotein signature, we employed the Likelihood-Ratio test (LRT) (22) and the Akaike information criterion (AIC) (23). Lastly, proportional-hazards assumptions were assessed with scaled Schönfeld residuals (Grambsch Therneau) (24) to ensure that the risk associated with a given score or parameter does not fluctuate significantly between early and late follow-up.

### Additional data

Additional details, data, and code are provided in the supplemental information, Supplementary Data 1 and Zenodo (doi: 10.5281/zenodo.21991905).

## Results

### Clinical and cardiopulmonary hemodynamic characteristics of the study cohort

The clinical characteristics of the analysed PAH patients are depicted in Table 1. Out of 101 patients, n=1 had to be excluded, because the NMR quality criteria were not met (Supplementary Note S1). The majority of patients were female and normal weight. At baseline, most of the patients had not started with any PAH therapy and about one third was anticoagulated, mostly due to atrial fibrillation (Table 2). The median survival time was 8 years (Fig. S2), but for those who died, the mean time to death was 3.6 years (Table 1). Pulmonary hemodynamics are detailed in Table 2. The remaining laboratory parameters are detailed in Table S1.

**Table 2:** Pulmonary hemodynamics and therapy of the PAH cohort at baseline (n=100) (mean±SD / median [quartiles]). The majority of patients were incident with no PAH medication or on PAH monotherapy for acral necrosis due to systemic sclerosis.

| Parameter | Value |
| --- | --- |
| Systolic pulmonary arterial pressure (mmHg) | 68 [45, 84] |
| Diastolic pulmonary arterial pressure (mmHg) | 24 [18, 33] |
| Mean pulmonary arterial pressure (mmHg) | 39 [28, 53] |
| Right atrial pressure (mmHg) | 6 [4, 10] |
| Pulmonary arterial wedge pressure (mmHg) | 8.8±3.2 |
| Central venous oxygen saturation (%) | 67.8 [60.5, 72.1] |
| Cardiac output (L/min) | 4.23 [3.47, 4.95] |
| Cardiac index (L/min/m<sup>2</sup>) | 2.34 [1.93, 2.83] |
| Arterio-venous difference of oxygen (%) | 4.85 [4.17, 5.66] |
| Pulmonary vascular resistance (WU) | 6.97 [4.17, 9.98] |
| Pulmonary vascular resistance index (WU*m<sup>2</sup>) | 13.5 [7.2, 18.3] |
| PVR / SVR – ratio | 0.38 [0.25, 0.57] |
| Total Pulmonary Resistance (WU) | 8.85 [6.25, 12.14] |

NMR-spectroscopy quantified 124 different metabolic and lipoprotein-related parameters in sufficient detail (<20% missing values) out of a total of 153 parameters. Creatinine was the only metabolite that was also available from routine clinical chemistry in all patients, showing a strong correlation with NMR-derived creatinine (r=0.85).

The heatmap revealed a cluster of 13 parameters, mainly composed of high-density lipoproteins of the HDL-1 and HDL-2 subclasses that were significantly associated with pulmonary hemodynamics as evident from the heatmap (Fig. 1A). PHIHDL was calculated from 12 of these parameters (see Supplementary Note S3 and Table S2).

**Fig. 1.**
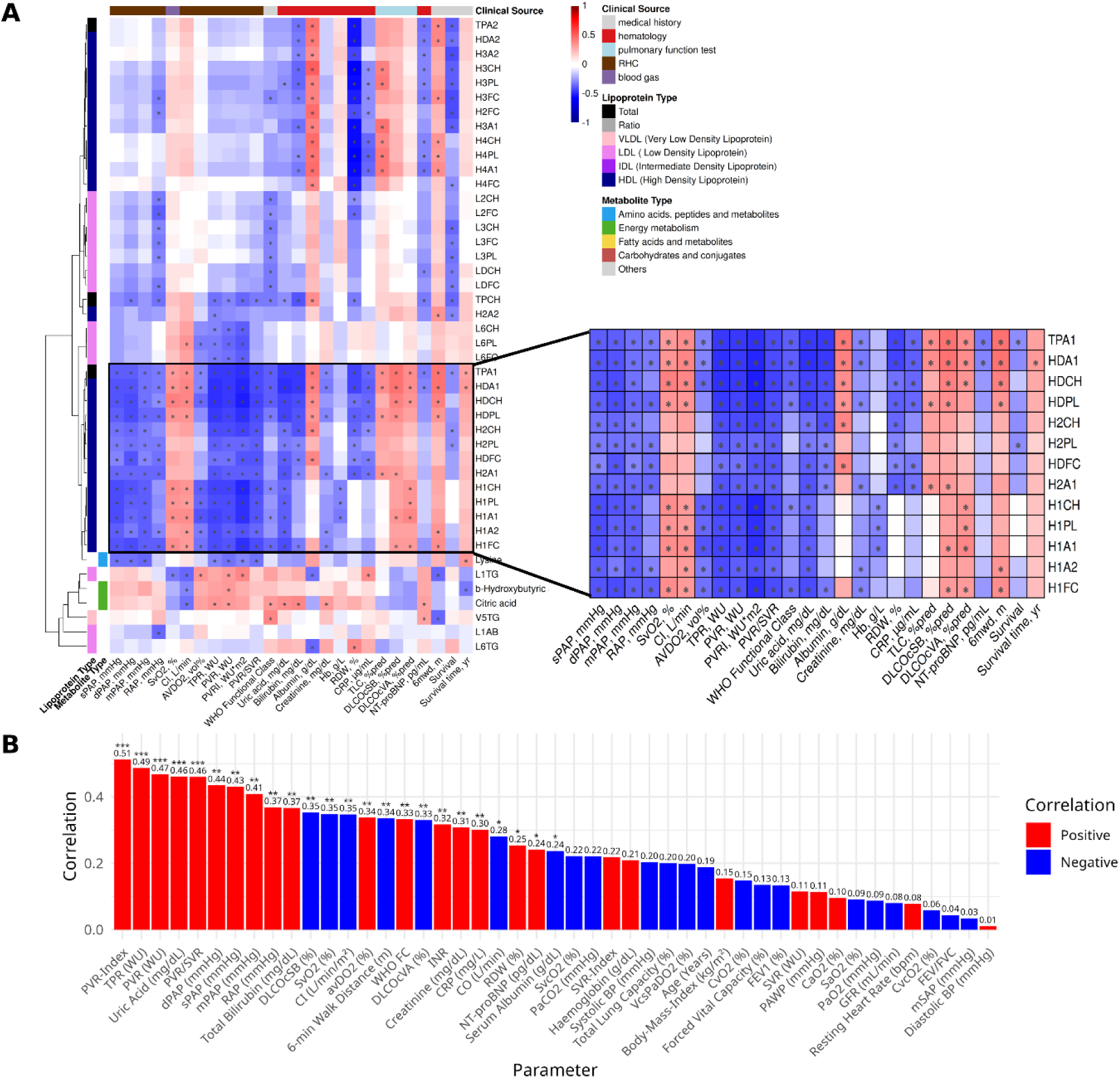
Spearman correlation heatmap of 44 NMR-derived metabolites and lipoprotein-related parameters vs. 26 clinical parameters. **(A)** Spearman correlation heatmap showing the associations between NMR-derived metabolites and clinical parameters. A cluster of HDL-related metabolites showing strong associations with pulmonary hemodynamics is highlighted and was used to derive **PHIHDL**. *, p < 0.05. **(B)** Spearman correlation of all clinical parameters with PHIHDL. Red, positive correlations, blue, negative correlations. *, p < 0.05. **, p < 0.01, ***, p < 0.0001.

As shown in Fig. 1B, PHIHDL was strongly associated with PVRI, representing the most direct measure of pulmonary vascular remodeling, and to many other pulmonary hemodynamic parameters, and also with uric acid, which is a well-known predictor of survival in PAH. In contrast, the association to systemic pressure, renal function and lung function was weak. A detailed description of the entire correlation analysis is presented in Supplementary Note S2.

The Kaplan Meier curves in Fig. S2A indicate the overall survival of the cohort including 1-year, 3-year, and 5-year survival rates, suggesting a substantial mortality risk despite inclusion of PH patients with mPAP between 21 and 24 and availability of all approved PAH drugs. Supplementary Table S3 shows the targeted PAH medication of the cohort at baseline and the final visit. Throughout the study, there was a considerable number of patients with mild pulmonary hemodynamics, who did not receive any PAH medication or just monotherapy, mostly originally prescribed for acral necrosis.

Survival was significantly associated with age and sex (Fig. S2B and C, Fig. S3). BMI was not significantly associated with survival or PHIHDL (r=0.15, p=0.13).

The performance of all prognostic scores is outlined in Fig. 3, with PHIHDL showing a significant association with survival, both with and without adjustment for age and sex. The prognostic role of other established markers is presented in Fig. S4. BNP, 6MWD and WHO-FC were significantly associated with survival, as well as creatinine, GFR, RDW, uric acid, and bilirubin, while hemodynamics like mPAP, RAP, and PVR were not significant (Fig. 4 and Fig. S5). This suggests that the prognostic function of PHIHDL goes beyond baseline pulmonary hemodynamics.

**Fig. 2.**
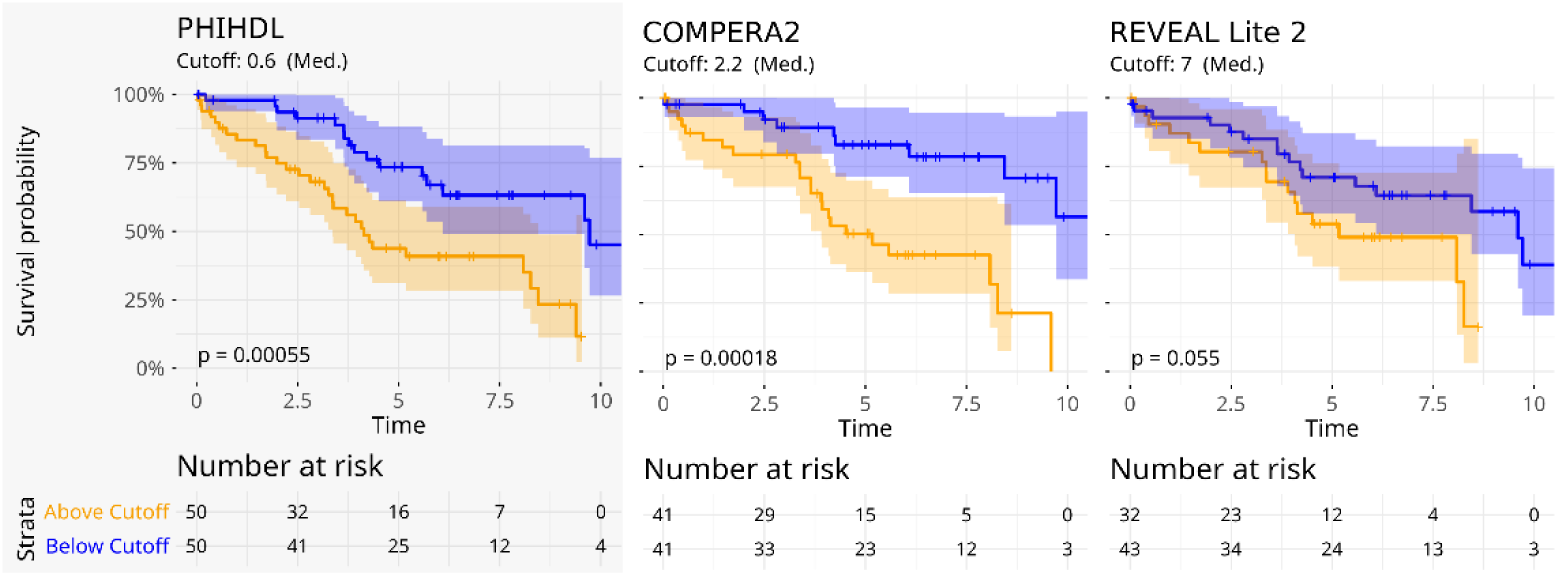
Univariate Kaplan-Meier survival analysis of PAH patients stratified by PHIHDL, COMPERA2, and REVEAL Lite 2 scores. Stratification using the median threshold for continuous scores.

**Fig. 3.**
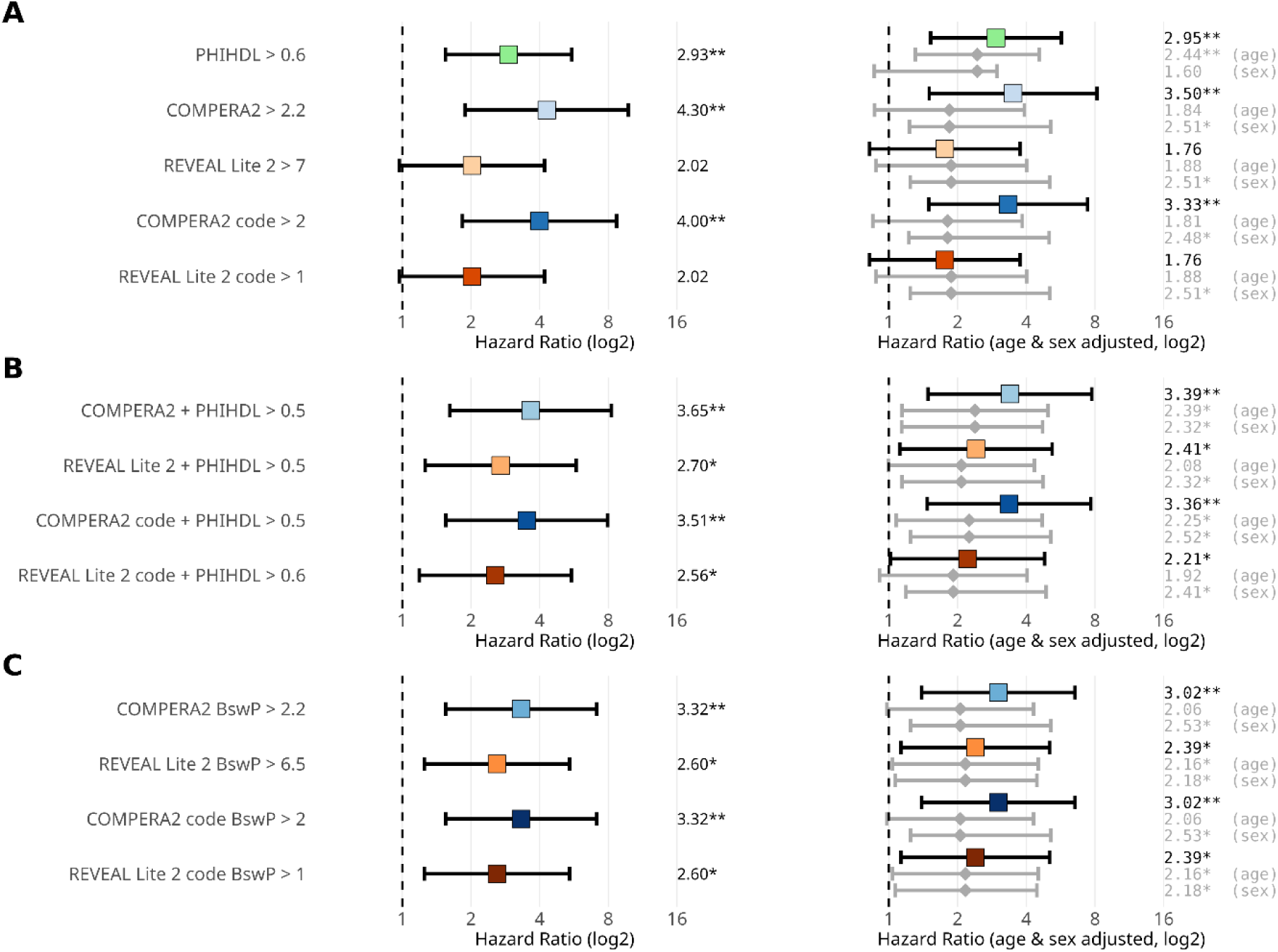
Cox-Proportional-Hazard for binarized scores. (A) Standalone scores. (B) Average-based score combinations. (C) Scores having BNP replaced by PHIHDL (BswP). Categorical “code” versions are based on the COMPERA2 four-strata and REVEAL Lite 2 three-strata groups and compared as “low + intermediate-low vs. intermediate-high + high” for COMPERA2 and “low vs. intermediate + high” for Reveal Lite 2.

**Fig. 4.**
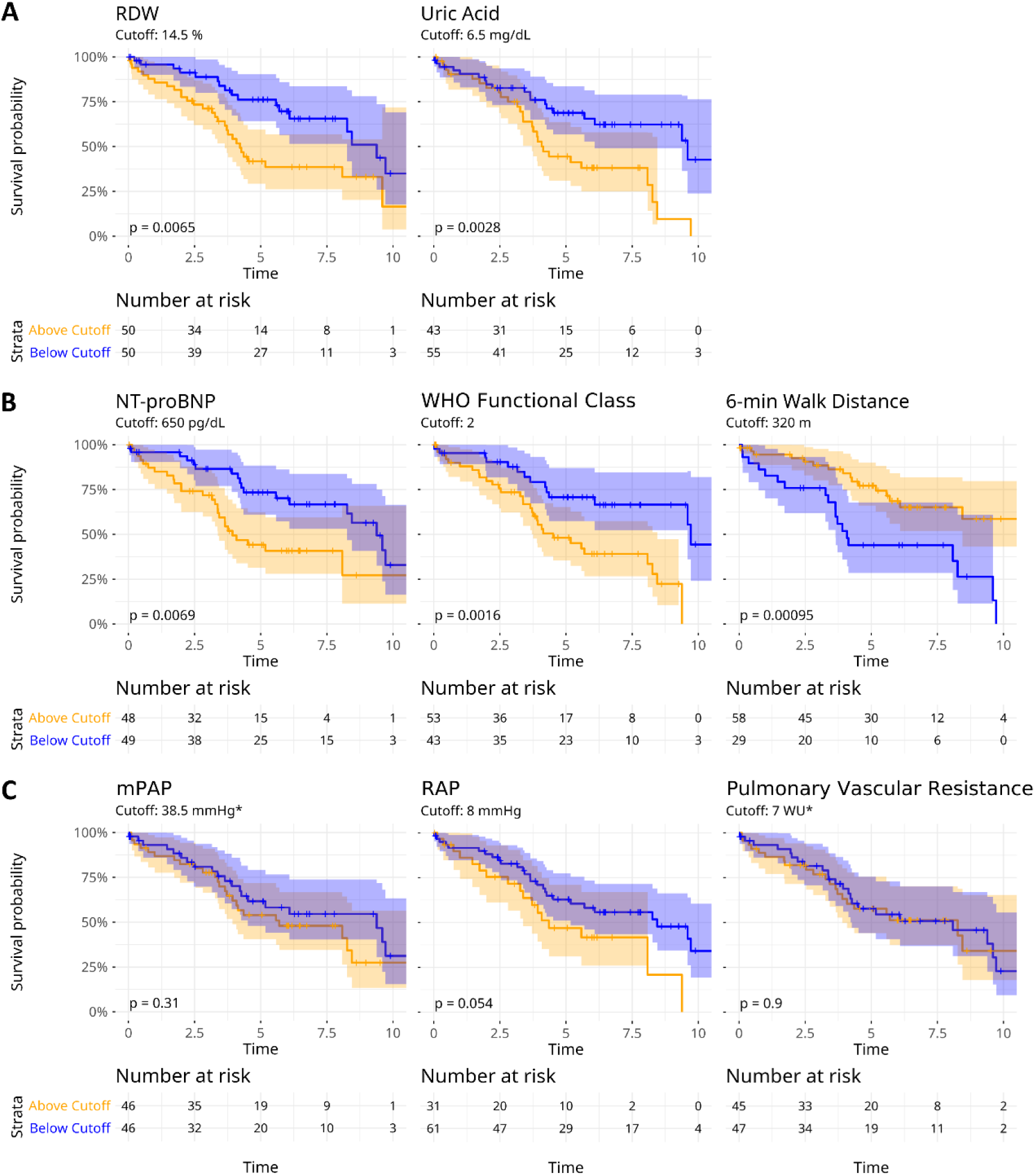
Univariate Kaplan-Meier curves of the cohort stratified by established predictors. **(A)** Potential biomarkers with prognostic value in PAH according to current literature. (**8,9**) **(B)** Parameters used for the calculation of COMPERA2. (**25**) **(C)** Hemodynamic parameters measured via RHC. Note that PVR, was not associated with prognosis. Thresholds were derived from literature unless marked with an asterisk *, which indicates the cohort median.

### PHIHDL predicts survival in PAH

We evaluated PHIHDL in comparison to established risk scores (COMPERA2, REVEAL Lite 2). To ensure comparability, all predictors were analyzed as dichotomized (binary) high vs. low (Fig. 2) by splitting at the cohort median. PHIHDL, COMPERA2 and REVEAL scores were positively associated with mortality (Fig. 2). In subsequent modeling, the effects of age and sex on the hazard ratios were assessed (Fig. 3).

As shown in Fig. 2, PHIHDL was highly significant, separating particularly well in the first three years, as compared e.g. to COMPERA 2 (PHIHDL vs. COMPERA2, 23% vs. 11% survival difference).

### PHIHDL complements existing scores

To assess whether PHIHDL provides information beyond established clinical risk tools, we first fitted an age-and sex-adjusted Cox model including both median-defined two-group predictors (COMPERA2 and PHIHDL). In this joint model, both COMPERA2 and PHIHDL remained independent significant predictors of survival (Fig. S6), indicating that each retains prognostic association when adjusted for the other (PHIHDL vs. COMPERA, HR 2.80 vs. 2.73; CI [1.16 – 6.75] vs. [1.09 – 6.83]; p-value 0.022 vs. 0.032). Next, in nested add-on comparisons within the same two-group framework, adding PHIHDL significantly improved model fit beyond both COMPERA2 and REVEAL Lite 2 (LRT p=0.007 / p = 0.001; Table 3) supporting its incremental prognostic value. Consistent with this, PHIHDL showed only moderate overlap with COMPERA2 (r=0.35) and an even weaker but still significant correlation with NT-proBNP (r=0.24, p=0.02).

**Table 3:** Incremental prognostic value of PHIHDL in age-and sex-adjusted Cox models. Predictors were dichotomized into high vs. low using the cohort median (median-based two-group approach). Likelihood-Ratio tests (LRT) and the Akaike information criterion (AIC) compare standalone vs. combined models (COMPERA2 vs. PHIHDL+COMPERA2 and REVEAL Lite 2 vs. PHIHDL+REVEAL Lite 2); negative ΔAIC indicates improved fit, with values below-2 suggesting meaningful improvement.

| Comparison | N<br>(events) | logLik<sub>base</sub> | logLik<sub>ext</sub> | p-value<sub>LRT</sub> | AIC<sub>base</sub> | AIC<sub>ext</sub> | ΔAIC |
| --- | --- | --- | --- | --- | --- | --- | --- |
| COMPERA2 vs.<br>COMPERA2 + PHIHDH | 82 (32) | -108.21 | -104.55 | 0.007 | 222.43 | 217.09 | -5.34 |
| REVEAL Lite 2 vs.<br>REVEAL Lite 2 + PHIHDH | 75 (32) | -109.90 | -104.91 | 0.001 | 227.10 | 218.33 | -8.77 |

Furthermore, to evaluate the prognostic utility of PHIHDL within established risk frameworks, we re-calculated the COMPERA2 and REVEAL Lite 2 scores using PHIHDL in place of NT-proBNP. The modified COMPERA2 and Reveal Lite 2 scores (COMPERA BswP and REVEAL Lite BswP) were equally predictive of survival as the original scores (Fig. 3C). The numerical loss in COMPERA2 and the numerical gain in REVEAL Lite 2 in the fully adjusted model were not significant.

Notably, when entered as a continuous, z-standardised variable, PHIHDL showed a hazard ratio of 1.61 (95 % CI 1.14-2.28) with an effect size comparable to that of the continuous COMPERA2 score (HR 1.86, 95% CI 1.23-2.82). In the long-term analysis, no meaningful departures from cox-proportional hazard assumptions were detected, implying that PHIHDL’s ability to predict outcomes is consistent throughout the study period. Detailed information including 95% confidence intervals is shown in Table S4. Cox models adjusting for only one covariate at a time (either age or sex) were also computed to visualize each covariate’s respective impact on survival (Fig. S7).

### Contribution of HDL-related measures to the prognostic function

To facilitate comparison with the study by Harbaum et al. (16), we evaluated the individual contributions of NMR-derived HDL subclasses and their measured constituents (cholesterol, phospholipids, free cholesterol, triglycerides, apolipoprotein (Apo) A1, ApoA2) to the prognostic performance of PHIHDL. HDL triglycerides did not contribute to prognostic performance, whereas all other components of HDL2 and HDL3 showed significant prognostic associations. Within HDL1, only free cholesterol was prognostic, and within HDL4, only ApoA1 was associated with outcome (Fig. 5).

**Fig. 5.**
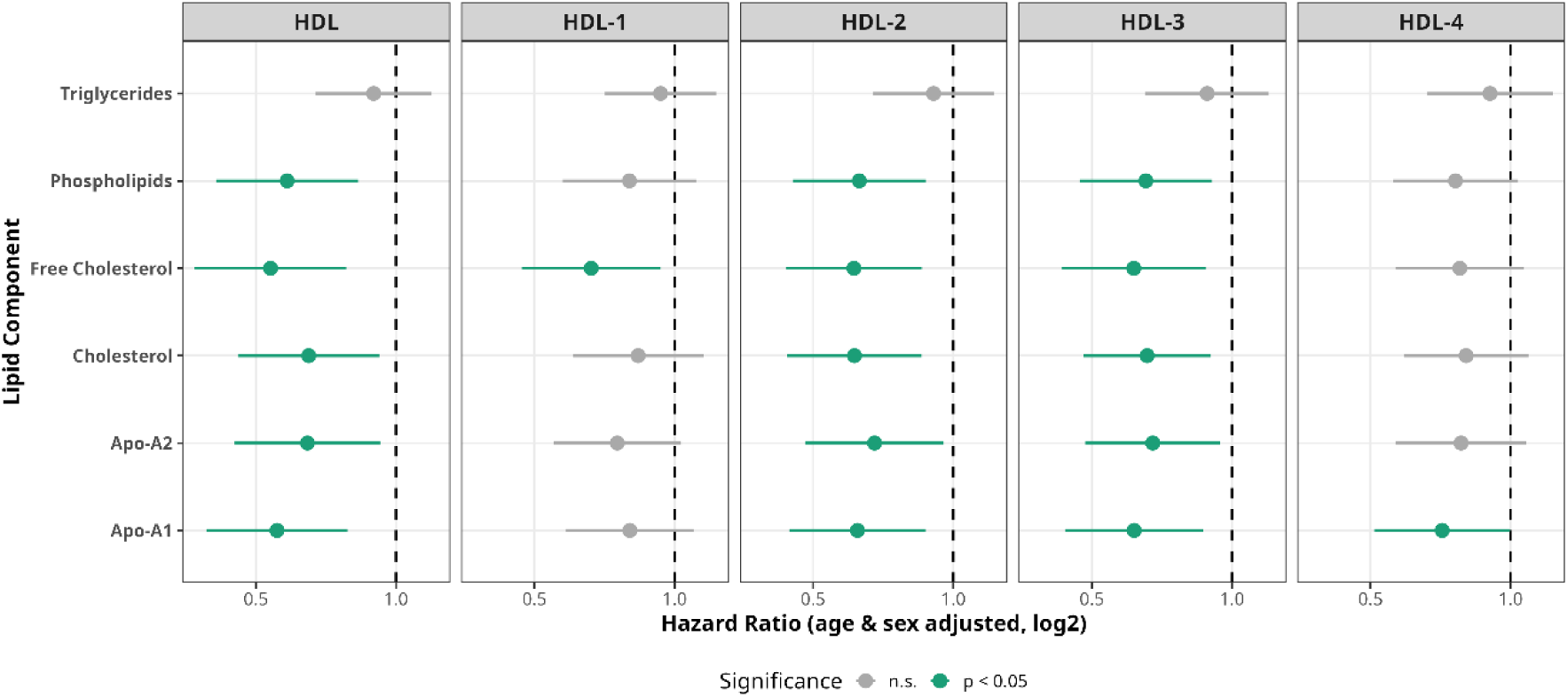
Survival analysis of HDL subclasses and their components. Age-and sex-adjusted hazard ratios for NMR-derived HDL measures within HDL1–HDL4. HR represents relative risk per 1 SD increase. Components of HDL2 and HDL3 showed the strongest associations with survival in PAH, whereas triglyceride-related measures were not significantly associated with survival in any HDL subclass.

## Discussion

In this study in PAH patients, comprehensive NMR-based profiling of circulating metabolites obtained during baseline right heart catheterisation identified large HDL subclasses (HDL1 and HDL2) as strongly associated with pulmonary but not systemic hemodynamics. From these subclasses, we derived PHIHDL, which was strongly associated with PVR, uric acid, WHO functional class, 6-minute walking distance, BNP, and DLCO. While PVR is the most direct measure of pulmonary vascular remodeling, the other factors are established markers of disease severity and prognosis in PAH (25). Importantly, PHIHDL was independently associated with long-term survival and, according to the Akaike information criterion, provided incremental prognostic information beyond the established clinical risk scores COMPERA2 and REVEAL2 lite.

Our findings are consistent with previous studies linking HDL biology to PAH outcomes. Heresi et al. reported that lower HDL-cholesterol levels were associated with poorer survival in PAH (15), and similar associations have been described in several retrospective cohorts (14,26,27) although not in the prospective French registry (28). A more recent study from Harbaum et al. (16), based on very similar NMR spectroscopy, compared to our study, also showed that low HDL levels were associated with poor survival, although small HDL fractions (HDL4) appeared to carry most of the prognostic information. We therefore analysed all the HDL-derived measures in our cohort, and found that all HDL2-and HDL3-derived measures, except triglycerides, were associated with mortality, whereas among HDL4-derived measures, only ApoA1 was prognostic (Fig. 5). This suggests that the prognostic information carried by HDL may depend on patient characteristics. We also reviewed the different subgroups included in our cohort (Fig. 6), although our study was not powered for specific subgroup analysis.

**Fig. 6.**
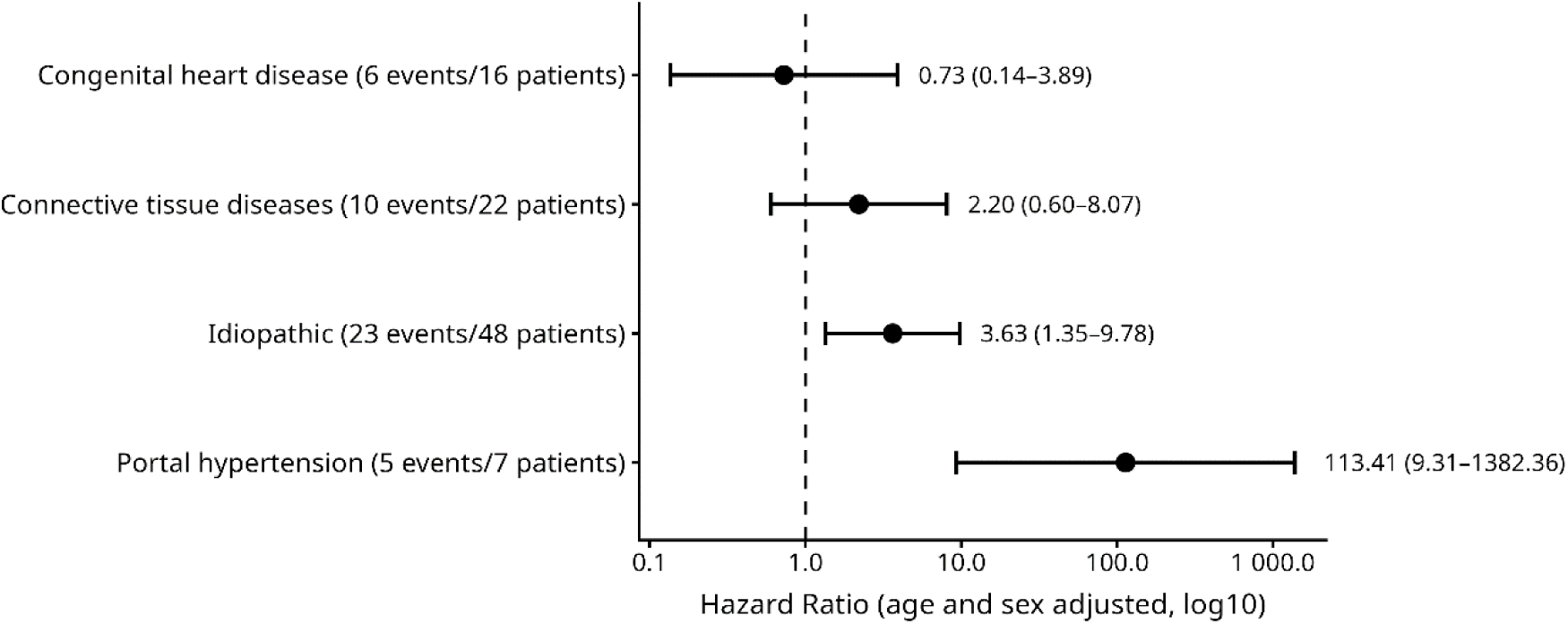
Prognostic function of PHIHDL by PAH subclass. Exploratory subgroup analysis applying a cox-proportional hazards model to showcase the interaction between elevated PHIHDL (>0.6) and RHC classification (p=0.005), adjusted for age and sex.

There were significant differences between PAH subgroups (p=0.005) and it seemed that PHIHDL was stronger associated to mortality in idiopathic PAH as compared to congenital heart disease. This indicates that HDL may be particularly sensitive for the molecular mechanisms involved in idiopathic PAH.

Our patients, as compared to the COMPERA2 and REVEAL Lite 2 patients, and the studies analysing the prognostic function of HDL (29) were comparable in terms of age, f/m ratio, and pulmonary hemodynamics, except that we also included patients with a mean pulmonary arterial pressure between 21 and 24 mmHg and that the vast majority of our patients were incident. Our baseline serum probes were from patients with mainly no PAH medication (58%), or just monotherapy (29%), mostly applied for acral necrosis due to systemic sclerosis (Table S3).

We derived PHIHDL by going for 12 HDL-1 and HDL-2 lipoprotein fractions, representing a strong band in our heatmap. The larger HDL fractions are known to be enriched with apo E1, in contrast to the smaller fractions. Indeed, apoE1 deficiency, in experimental models, was strongly associated to pulmonary hypertension (30–32), which may represent a mechanistic link to our findings.

Although some of the parameters used for PHIHDL could have been used as standalone markers, our approach of integrating a broader panel of particles may have enhanced the score’s overall stability and prognostic separation power. We chose this most conservative approach to avoid optimization bias. Technically, PHIHDL was easy to assess and could provide a fast and low-cost parameter for clinical routine.

Current PAH risk stratification relies mainly on clinical parameters incorporated into scores such as COMPERA2 and REVEAL Lite 2 (2,17,25). Both scores include WHO functional class, 6-minute walking distance, and BNP, causing strong associations between the scores and their components (Fig. S8). After correction for age and sex, only COMPERA2 and PHIHDL remained significant prognostic scores. Additional formal statistical testing showed that PHIHDL was independent of COMPERA2 and vice versa (Fig. S6) and that combination of PHIHDL with COMPERA2 and REVEAL Lite 2 scores, according to the Akaike information score, added prognostic information to both COMPERA2 and REVEAL (Table 3). This suggests that PHIHDL captures biological information beyond established clinical risk scores.

### Clinical implications

The implementation of PHIHDL into clinical practice could leverage existing high-throughput, NMR-based lipid-profiling platforms. This approach offers the potential for a highly standardized and automated method of risk assessment, as highlighted in Fig. 7. The approach is based on our median values for point assignment in existing prognostic frameworks. Similar to COMPERA2, Reveal Lite 2 uses four groups for point assignment but awards-1.5 points below, and 1.5 points above the threshold in this version.

**Fig. 7.**
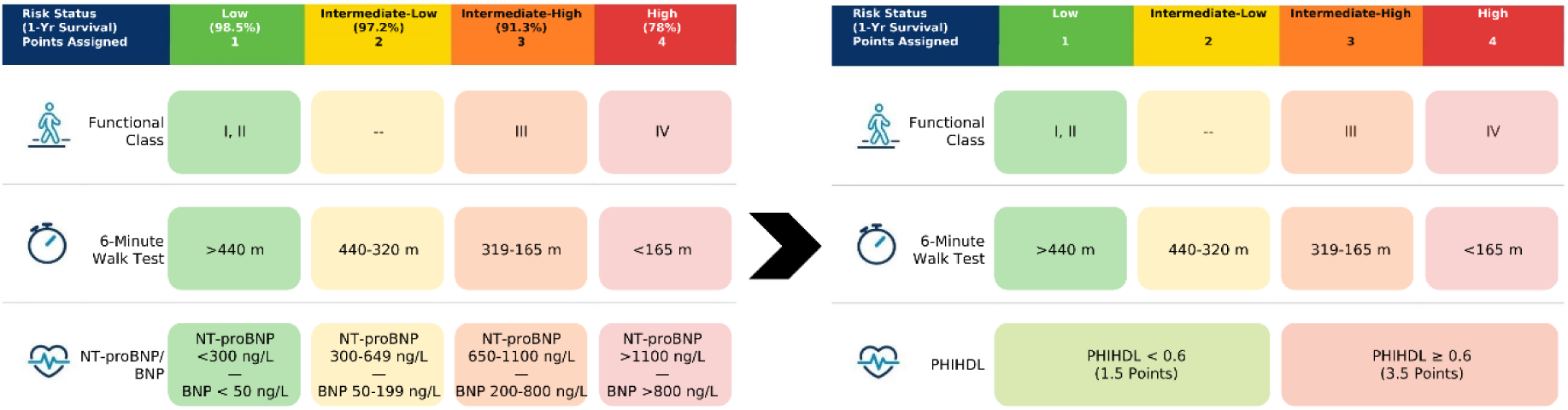
Conceptual adaptation of the COMPERA2 risk score incorporating PHIHDL to evaluate its prognostic utility within established frameworks. Due to sample size constraints, PHIHDL was dichotomized into low-and high-risk categories. The assigned points reflect the mean NT-proBNP contribution in the original COMPERA2 model. Graphic adapted from Dardi et al. (**25**)

### Potential molecular mechanism behind PHIHDL’s prognostic function

Patients with PAH exhibit profound metabolic/lipoprotein alterations, including changes in lipid metabolism, (10,13,33) and kynurenine metabolism (34,35), and circulating HDL measures (14–16,26), while insulin resistance remains a controversial topic (10,11).

Notably, PHIHDL was derived based on associations with pulmonary hemodynamics but showed stronger associations with survival than baseline PVR, suggesting that it captures additional aspects of disease biology, while baseline PVR loses any prognostic function during PVR-lowering therapies.

In coronary artery disease patients, elevated HDL-cholesterol is independently associated with event-free and overall survival (36). The most widely discussed mechanism underlying this association is HDL-mediated reverse cholesterol transport, whereby HDL promotes the removal of excess and oxidized cholesterol species, such as 7-ketocholesterol, from peripheral tissues and facilitates their delivery to the liver for biliary excretion (37).

Beyond lipid transport, HDL reduces oxidative stress and may reduce pulmonary artery smooth muscle cell proliferation, processes central to pulmonary vascular remodeling (38,39). In addition, circulating HDL levels are associated with flow-mediated vasodilation (40,41) and coronary vasoconstrictor responses (42), and infusion of recombinant HDL particles in individuals with isolated low HDL levels normalized impaired forearm blood flow (43). In case of PAH, high levels of circulating fatty acids are associated with a poor prognosis (13). HDL has a considerable capacity to serve as a sink for oxidized lipids (44), which might ameliorate the detrimental fatty acid effects in PAH.

Notably, PHIHDL is primarily composed of measures derived from larger HDL particles, which are enriched in apoE (45–47). Experimental studies have demonstrated that ApoE deficiency promotes pulmonary hypertension and pulmonary vascular remodeling, particularly under high-fat dietary conditions (31). Mechanistic studies further suggest that apoE modulates signalling through bone morphogenetic protein receptor type 2 (BMPR2), (30) a central pathway in PAH pathobiology (3). Although ApoE cannot be quantified by the NMR platform used in this study, enrichment of larger HDL particles with apoE provides a potential biological link that may contribute to the prognostic associations observed with PHIHDL. It might also explain why its prognostic function was superior in idiopathic as compared to congenital heart disease-associated PAH.

### Strengths and limitations Strengths

Our cohort consisted of PAH patients from a prospective registry who all received a right heart catheter investigation by the same experienced team. All patients were carefully checked for any causes of pulmonary hypertension other than PAH, using a large number of clinical investigations and the majority of the patients were incident. Blood samples were collected during the baseline right heart catheterization, without the need for fasting or dietary restrictions. Patients did not require any diet or overnight starve before blood was drawn for PHIHDL assessment and PHIHDL could be determined in all patients. This makes it an attractive option for use in clinical algorithms.

### Limitations

Firstly, this was a monocentric study with strict inclusion criteria, making the patient cohort relatively small. However, the follow-up was long and complete. The sample size also precluded more complex prognostic model optimisation, which would require independent validation. Secondly, PHIHDL was derived from unsupervised correlation analysis rather than mechanistic selection. Nevertheless, this approach is consistent with previous studies and may point to novel mechanistic links for pulmonary arterial remodeling. Third, only baseline measurements were available, therefore, the value of PHIHDL for longitudinal monitoring or treatment guidance remains unknown. Fourth, the NMR method used in this study had limitations related to the sensitivity for metabolites other than lipoproteins. Such metabolites, e.g. fatty acids, could add further important diagnostic and prognostic information. Finally, survival analysis stratified by therapy were not feasible due to multiple therapeutic changes and many combination therapies, limiting statistical power.

## Conclusion

PHIHDL represents a pulmonary hemodynamics-based metabolomic parameter that is independently associated with long-term survival and may complement established clinical risk scores for risk stratification in PAH and may be associated with the pathologic mechanisms of idiopathic PAH.

### Outlook

PHIHDL, primarily derived from HDL1-and HDL2-related measures, is associated with pulmonary hemodynamics and provides independent prognostic information in PAH. Its biological basis remains to be determined but may involve HDL-mediated anti-inflammatory, antioxidative, endothelial-protective, and BMPR2-related pathways. Validation in larger cohorts and mechanistic studies is warranted.

### Short summary

PHIHDL, a metabolomic score from large HDL particles, was derived based on its association with pulmonary hemodynamics and independently predicts survival in patients with pulmonary arterial hypertension (PAH), in particular idiopathic PAH. Its biological basis may involve BMPR2-related pathways. PHIHDL may represent a potential laboratory-based component for future PAH risk stratification.

### Author contributions

Conceptualisation, HO, AO, NB; Data curation, SP, NB, VF, GK, HO, AO; Formal analysis, SP, NB, HB, HH, TM; Funding acquisition, HO, AO; Investigation, VF, HH, TM, GK; Methodology, SP, NB, HH, TM, HO, AO; Project administration, SP, NB, UB, HO, AO; Resources, SP, NB, HB, HH, TM; Software, SP, NB, HB; Supervision, NB, HH, UB, HO, AO; Validation, SP, NB, VF, HB, HH, TM, GM, CN, GK, HO, AO; Visualisation, SP, HB; Writing – original draft, SP, TM, GM, GAH, HO, AO; Writing – Review & Editing, all authors;

## Data Availability

All data produced are available online at https://doi.org/10.5281/zenodo.21991904

## Acknowledgements

We are very grateful for the excellent technical assistance from Mag. Dr. Daniela Kleinschek. The samples/data used for this project have been provided by Biobank Graz of the Medical University of Graz, Austria: Cohort GRAPH-M. T.M. thanks the Center for Medical Research, Medical University of Graz, Graz, Austria for laboratory access. T.M. is grateful to the Austrian Science Fund (FWF) for excellence cluster 10.55776/COE14, Grants DOI 10.55776/P28854, 10.55776/I3792, 10.55776/DOC130, and 10.55776/W1226, the Austrian Research Promotion Agency (FFG) grants 864690 and 870454; the Integrative Metabolism Research Center Graz; the Austrian Infrastructure Program 2016/2017; the Styrian Government (MetAGE, Zukunftsfonds, doc.fund program); the City of Graz (MetAGE, doc.fund); and BioTechMed-Graz (flagship project). This project was funded in part by the FFG and the European Union (EFRE) under grant 912192. For open access purposes, the author has applied a CC BY public copyright license to any author accepted manuscript version arising from this submission.

## Abbreviations

(NT-pro)BNP: (N-terminal pro-) b-type natriuretic peptide
6MWD: six minute walking distance
AIC: Akaike Information Criterion
Apo: apolipoprotein
AVDO2: arterio-venous difference in oxygen content
BMI: body mass index
CaO2: arterial oxygen content
CI: cardiac index
CO: cardiac output
COPD: chronic obstructive pulmonary disease
CRP: C-reactive protein
CvcO2: vena cava superior oxygen content
CvO2: venous oxygen content
DLCOcSB: diffusion capacity for carbon monoxide (single breath)
DLCOcVA: diffusion capacity for carbon monoxide (alveolar volume)
dPAP: diastolic pulmonary artery pressure
dSAP: diastolic systemic artery pressure
FEV/FVC: FEV1 over FVC
FEV1: forced expiratory volume 1s
FVC: forced vital capacity
GFR: glomerular filtration rate
Hb: hemoglobin
HDL: high density lipoproteins
HR: hazard ratio
HMDB: Human Metabolome Database
INR: international normalised ratio
KM: Kaplan-Meier
mmHg: millimeter of mercury
LRT: Likelihood-ratio test
mPAP: mean pulmonary artery pressure
mSAP: mean systolic artery pressure
NMR: nuclear magnetic resonance
PaCO2: arterial partial pressure of carbon dioxide
PAH: pulmonary arterial hypertension
PaO2: arterial partial pressure of oxygen
PAWP: pulmonary artery wedge pressure
PH: pulmonary hypertension
PHIHDL: pulmonary hypertension index from high density lipoproteins
PVR: pulmonary vascular resistance
PVRI: pulmonary vascular resistance index
RAP: right atrial pressure
RDW: red-cell distribution width
RHC: right heart catheterization
SaO2: arterial oxygen saturation
SMWM: small molecular weight metabolites
sPAP: systolic pulmonary artery pressure
sSAP: systolic systemic artery pressure
SVO2: mixed venous oxygen saturation
SVR: systemic vascular resistance
SVRI: systemic vascular resistance index
TLC: total lung capacity
WHO FC: world health organization functional class
WU: Wood units

## Supplementary Notes

### S1) Data Preprocessing and Quality Control

To ensure analytical and biological integrity, metabolic parameters underwent a multi-stage quality control pipeline. Initial screening for biological plausibility was conducted by cross-referencing measured concentrations with established physiological reference ranges from the Human Metabolome Database (HMDB) (48) and the linear quantification ranges specified by Bruker. Univariate outlier detection was performed through visual inspection of distribution plots and supported by the median absolute deviation (MAD) (49) method; however, no parameters exceeded the predetermined significance thresholds.

Multivariate outlier detection was executed via principal component analysis (PCA) on the total parameter set and thematic subsets (clinical data, lipoproteins, and small-molecular-weight metabolites), identified one outlier sample which was subsequently excluded. (50)

While lipoproteins showcased virtually no missing values (except for 5 parameters belonging to VLDL subclasses or IDL triglycerides), a missingness threshold of 20% was utilized to preserve small-molecular-weight metabolites. This way we preserved these biologically relevant features that might otherwise be excluded due to concentrations approaching the analytical limit of detection (LOD) or limit of quantification (LOQ) in specific sub-groups of patients. The clinical parameter subset was mostly complete, except eight samples retrieved 3–6 months post-RHC in cases with no or unchanged PAH therapy, and five baseline examinations without pulmonary function testing. Other than that, missingness was constrained mostly to few isolated values occurring at random within routine clinical laboratory testing. Estimated glomerular filtration rate (eGFR) was derived from creatinine clearance, with adjustments made for patient age, sex, and body surface area (BSA).

Since NMR measurements below the limit of detection are not truly missing, relevant values were imputed. For small-molecular-weight metabolites, this was done by using half of the provided LODs as the imputed value. Since lipoprotein measurements are harder to quantify, empirical limits were estimated utilizing half the 2.5% quartile of observed non-zero values.

Following quality control validation and imputation, low-molecular-weight metabolites were log_10_-transformed to stabilize variance and mitigate right-skewness, adhering to established standards for the statistical analysis of metabolomics data. (51)

### S2) Identification of lipoprotein signatures in pulmonary arterial hypertension

We performed a comprehensive correlation analysis, computing pairwise Spearman coefficients between 124 metabolic particle concentrations and a panel of 50 established clinical and physiological parameters. Due to spatial constraints, metabolites were excluded if they were not significantly correlated with any clinical attributes, resulting in the 92-metabolite by 50-clinical heatmap detailed in Fig. S9A (including RHC-derived hemodynamics, blood gas analysis, hematology, and pulmonary function tests).

To focus on the most meaningful relationships, we then applied a data-driven filtering strategy. We defined valid associations as those meeting a significance threshold of p<0.05 after adjustment via Benjamini-Hochberg and with a correlation coefficient of r>0.1. This conservative correlation threshold was selected to capture even mild associations - consistent with Cohen’s benchmarks for small effect sizes - ensuring that potentially relevant biological signals were not prematurely discarded. (52) Subsequently, using a scree-like heuristic (21), where the cutoff was set at the first major inflection point (the ‘knee’), we established an optimal inclusion threshold requiring each metabolic parameter to have at least three significant correlations to be retained (Fig. S1A). Clinical parameters were filtered in a similar fashion, requiring a minimum of one significant correlation with metabolic parameters (Fig. S1B). This approach aimed to balance the removal of weakly associated parameters against significant data loss and resulted in a 44-metabolite by 26-clinical parameter heatmap.

Hierarchical clustering of the filtered data (using the complete linkage method) revealed a distinct cluster of lipoproteins that was strongly correlated with key hemodynamic parameters (Fig. 1). The high degree of co-correlation among parameters within this cluster, especially those related to ApoA-I and HDL, suggested that a composite score could serve as a more robust indicator of disease severity and possibly of prognosis. Due to broad overlap between TPA1 and HDA1, TPA1 was ignored in subsequent score calculations with further details being presented in Supplementary Note S3.

Consequently, we defined a novel “Pulmonary Hypertension Index from High Density Lipoproteins” (PHIHDL) score by averaging the individually MinMax-scaled (0–1) values of 12 selected lipoprotein particles from this cluster. These parameters were complete, i.e. there were no missing values across patients. We selected MinMax scaling over z-score or IQR standardization to generate a bounded score (0–1) that is intuitive for clinical interpretation. The choice of scaling method had no impact on performance due to our subsequent stratification approach (dichotomization by median), which ensures stability regardless of the underlying scale. While PHIHDL showed a significant association with survival, a sensitivity analysis confirmed that the cluster’s structure was not primarily driven by survival (Fig. S9B).

Finally, to understand the clinical relevance of this novel score, we examined its association with established parameters. PHIHDL was strongly correlated with critical pulmonary hemodynamic measures (PVR, sPAP, SvO₂, mPAP) as expected from the selection process for its components. However it was also strongly associated with e.g. uric acid, which is a well-known PAH prognosticator (Fig. S8A).

### S3) Sensitivity analyses addressing overlapping components

The identified cluster comprised 13 lipoprotein parameters, several of which are aggregate measures that overlap with their discrete constituents (e.g., HDA1 overlaps with H1A1 and H2A1). TPA1 and HDA1 in are biologically almost identical, as Apo-A1 is the main protein in HDL and >90% of serum Apo-A1 is present in the HDL subclass. (53,54) This overlap can induce implicit re-weighting and potential double-counting. We therefore a) treated TPA1 and HDA1 as equivalent (removing TPA1, as it is less specific) and b) deconstructed PHIHDL to reveal the underlying subclass-specific components. All components are outlined in their original (non-inverted) version for ease of interpretation.

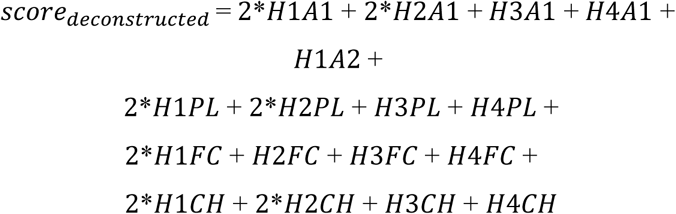

This listing of components reveals that within PHIHDL, triglyceride components and Apo-A2 are under-represented (or absent), whereas HDL-1 and HDL-2 are weighted more heavily due to their occurrence in both aggregate and discrete measures which make up PHIHDL (e.g., H1A1 enters both as a constituent of HDA1 and individually as H1A1 = 2 * H1A1).

While this addresses the problem of overlapping attributes, we decided to assess the robustness of PHIHDL by conducting the following sensitivity analyses:

1) Aggregate-Level Model: We restricted the analysis to aggregate class-level parameters. This determines if the predictive signal remains robust when using standard, lower-resolution clinical metrics.

a. Parameters: HDA1, HDPL, HDFC, HDCH
2) Subclass-Resolved Model: We utilized only the discrete subclass-specific parameters. This approach isolates the high-resolution signal. Note that this configuration inherently omits HDL-3 and HDL-4 constituents, as those sub-classes only contributed to the original score via the aggregate totals mentioned in approach 1.

a. Parameters: H1A1, H2A1, H1A2, H1PL, H2PL, H1FC, H1CH, H2CH
3) Unweighted Model: We evaluated the deconstructed score by assigning unit weights to all unique components. By removing redundant variables and eliminating weighting (coefficients), we tested whether double counting improves the metabolic signal.
a. Parameters: H1A1, H2A1, H3A1, H4A1, H1A2, H1PL, H2PL, H3PL, H4PL, H1FC, H2FC, H3FC, H4FC, H1CH, H2CH, H3CH, H4CH

**Approach 1**, while still significantly associated with survival, reduced HR compared to PHIHDL (HR 2.40 vs. 2.95). This indicates that within-HDL subclass structure (HDL-1 & 2 vs. HDL-3 & 4) carries prognostic information that is diluted, when averaged. However, this score may have value in combination with COMPERA2 code and Reveal Lite 2, increasing the HR vs. the combination with PHIHDL (HR 4.20 vs. 3.36 for COMPERA2 code, HR 2.60 vs. 2.41 for Reveal Lite 2).

**Approach 2**, very similar to approach 1, preserved statistical significance. For the combination of PHIHDL with Reveal Lite 2, this slightly improved the hazard ratio gained in approach 1 vs. PHIHDL (HR 2.80 vs. 2.41).

**Approach 3** results in plummeting prognostic performance vs. PHIHDL (HR 2.13 vs. 2.96) but improves prognostic performance in combination with COMPERA2 and COMPERA2 code vs. PHIHDL (HR 4.08 vs. 3.50, and HR 4.77 vs. 3.33). The former can be attributed to a) the reduced weight of HDL-1 and HDL-2 and b) the increased influence of measurement error and model-related uncertainty introduced during spectral mixture analysis and PLS-based quantification by the Bruker algorithm (noise) at the sub-class level, specifically observed for free cholesterols (see Supplementary Data 1).

As Harbaum et. al., found the strongest prognostic information in the small HDL particles, particularly HDL4, (16) we compared the prognostic information contained in HDL1+HDL2 vs. HDL3+HDL4 in particular.

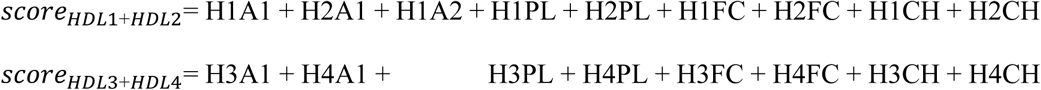

The comparison of the prognostic performances indicates that HDL1+2 score is roughly equivalent to PHIHDL (HR 2.77 vs. 2.96), while HDL3+4 score loses its predictive power (HR 1.49, NS). On the other hand, in combination with COMPERA2 code, HDL3+4 improved the prognostic power (HR 4.14 vs. 3.33).

Detailed Cox model results are also shown in Supplementary Table S5, indicating that PHIHDL performs best while alternative scores are also dependent on sex as a covariate.

### S4) Use of optimization algorithms for group separation

We further tested an optimized thresholding method for survival stratification. While this approach showed significant prognostic power (Fig. S10), it tended to create small, extreme-risk groups. Given the lack of a validation cohort, we adopted the more conservative strategy of using the median as the universal cutoff for all scores to optimize for robustness.

### S5) Evaluating the prognostic utility of PHIHDL within established risk frameworks

To evaluate the predictive utility of PHIHDL for survival, we assessed its integration into established risk models. Although PHIHDL showed only a weak correlation with NT-pro-BNP (r=0.24), its strong association with pulmonary hemodynamics suggests that it captures a distinct pathophysiological dimension. To evaluate this independently, we calculated modified versions of both scores using PHIHDL values (scaled 0–3 points) in place of the NT-proBNP component of COMPERA and REVEAL scores. Due to the limited sample size, we assessed the potential improvement in risk stratification using a dichotomized approach, as shown in Fig. 7. For combination models, all scores were MinMax scaled to a 0–1 range to ensure comparable weighting.

### S6) Combination with existing biomarkers

To explore the potential for improved prognostication, we investigated whether a nonlinear combination of existing survival-related parameters and PHIHDL could enhance the predictive accuracy of COMPERA2 and REVEAL lite 2. A two-dimensional point cloud visualization (Fig. S11) was used to examine relationships between PHIHDL and various parameters, color-marked by 3-year survival status. This confirmed that higher PHIHDL was associated with 3-yr mortality and suggested that particularly the combination of high PHIHDL with low DLCO was strongly associated with high 3yr-mortality (Fig. S11). The same was true for the combination of high PHIHDL with low serum albumin and low SvO2.

## Supplementary Tables

**Table S1:** Overview of the analysed PAH patients. N=100, Mean±SD and Median [quartiles].

| Parameter | Value |
| --- | --- |
| **Pulmonary hemodynamics from RHC** |  |
| Resting heart rate (bpm) | 72±13 |
| Mean systemic arterial pressure (mmHg) | 83±11 |
| Mean pulmonary arterial pressure (mmHg) | 39 [28, 52] |
| Pulmonary arterial wedge pressure (mmHg) | 8.8±3.2 |
| Right atrial pressure (mmHg) | 6 [4, 10] |
| Cardiac output (L/min) | 4.23 [3.47, 4.95] |
| Systemic vascular resistance (WU) | 17.51 [14.25, 22.30] |
| Pulmonary vascular resistance (WU) | 6.97 [4.17, 9.98] |
| Total pulmonary resistance (WU) | 8.85 [6.25, 12.14] |
| Cardiac index (L/min/m<sup>2</sup>) | 2.34 [1.93, 2.83] |
| Systemic vascular resistance index | 31.2 [24.1, 38.7] |
| Pulmonary vascular resistance index | 13.6 [7.2, 18.3] |
| Systolic pulmonary artery pressure (mmHg) | 68 [45, 84] |
| Diastolic pulmonary artery pressure (mmHg) | 24 [18, 33] |
| Arterial oxygen saturation (%) | 94.4 [92.3, 96.1] |
| Central venous oxygen saturation (%) | 67.8 [60.5, 72.1] |
| Total arterial oxygen content (%) | 17.5 [16.1, 19.0] |
| Total venous oxygen content (%) | 12.7 [11.1, 13.9] |
| Arterio-venous difference of oxygen (%) | 4.85 [4.17, 5.66] |
| PVR over SVR | 0.38 [0.25, 0.57] |
| **Blood count and clinical chemistry** |  |
| NT-proBNP (pg/dL) | 645 [174, 1893] |
| Creatinine (mg/dL) | 0.98 [0.80, 1.17] |
| Glomerular filtration rate (mL/min) | 69.9±23.3 |
| Uric acid (mg/dL) | 6.4 [5.1, 7.7] |
| Total bilirubin (mg/dL) | 0.7 [0.4, 1.0] |
| C-reactive protein (mg/L) | 2.8 [1.4, 5.8] |
| Red cell distribution width (%) | 14.6 [13.5, 15.9] |
| Haemoglobin (g/dL) | 14.0±2.1 |
| **Pulmonary function test** |  |
| Forced vital capacity (% predicted) | 87.3±19.2 |
| Forced expiratory volume 1s (% predicted) | 80.2±19.0 |
| FEV1/FVC | 76.5±10.9 |
| Total lung capacity (% predicted) | 95.0 [81.6, 107.0] |
| CO diffusing capacity related to body surface, Hb corr. (% predicted) | 57.7±22.3 |
| KCO diffusing capacity related to alveolar volume, Hb corr. (% predicted) | 68.2±25.3 |

**Table S2:** Parameters composing PHIHDL and their individual components (mean±SD). (Terminology according to Bruker) Stars * denote that the underlying data is not normally distributed but is presented using mean±SD to facilitate the understanding of the lipoprotein landscape. N=100.

| Parameter | Abbreviation | Mean±SD | Discrete Components |
| --- | --- | --- | --- |
| HDL Apo-A1 | HDA1 | 147.1±28.5 | H1A1 + H2A1 + H3A1 + H4A1 |
| HDL cholesterol | HDCH | 57.2±14.4 | H1CH + H2CH + H3CH + H4CH |
| HDL phospholipids | HDPL | 84.3±16.7 | H1PL + H2PL + H3PL + H4PL |
| HDL free cholesterol | HDFC | 15.3±4.7 | H1FC + H2FC + H3FC + H4FC |
| HDL-1 Apo-A1 (mg/dL) | H1A1 | 30.3±17.9* | - |
| HDL-2 Apo-A1 (mg/dL) | H2A1 | 20.8±4.4 | - |
| HDL-1 Apo-A2 (mg/dL) | H1A2 | 3.3±1.6* | - |
| HDL-1 cholesterol (mg/dL) | H1CH | 19.6±10.7 | - |
| HDL-2 cholesterol (mg/dL) | H2CH | 9.8±2.3 | - |
| HDL-1 phospholipids (mg/dL) | H1PL | 25.5±12.1 | - |
| HDL-2 phospholipids (mg/dL) | H2PL | 15.7±3.4 | - |
| HDL-1 free cholesterol (mg/dL) | H1FC | 5.7±2.4* | - |

**Table S3:**
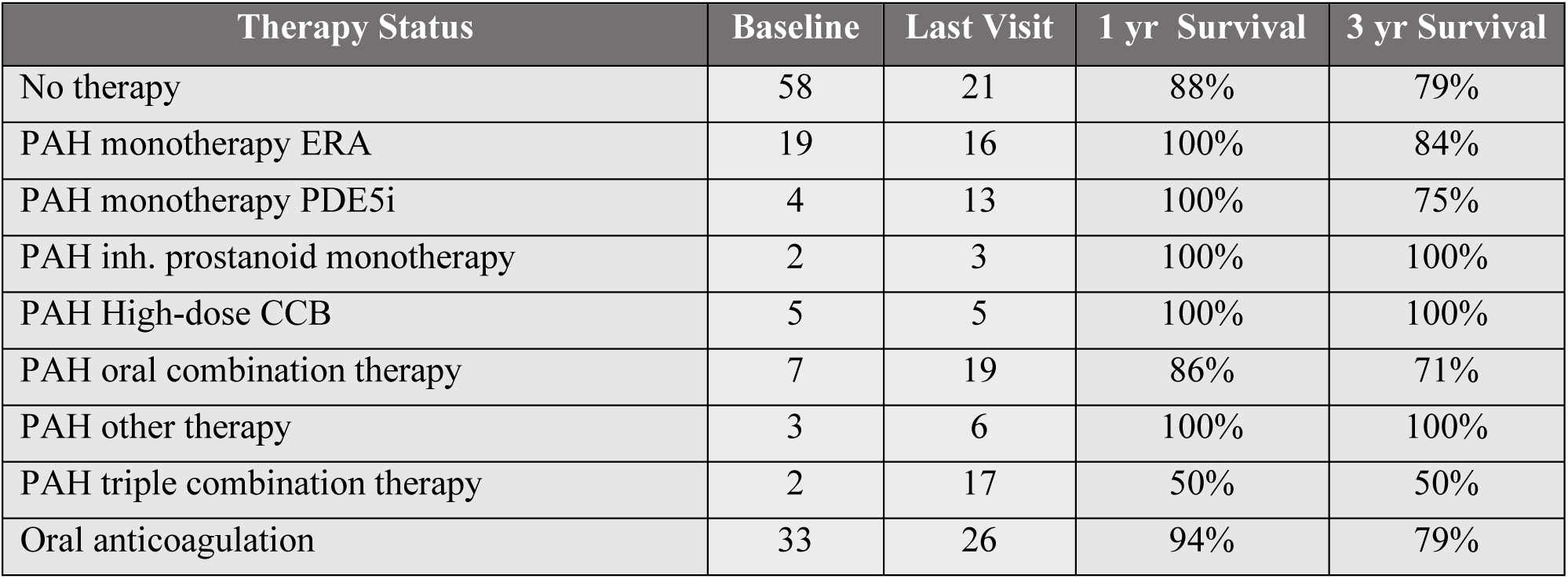
Distribution of PAH therapy status over time. Data are represented as absolute counts (N) for the cohort at baseline and last clinical follow-up. 1-and 3-year survival is given for each therapy status at baseline.

**Table S4:** Cox proportional-hazard models for all relevant scores. Scores have been tested standalone, in combination with PHIHDL and by evaluating PHIHDL inside the COMPERA2 and REVEAL Lite 2 frameworks.

| Score | N<br>(number of events) | HR | 95% CI | p-value |
| --- | --- | --- | --- | --- |
| PHIHDL > 0.6 | 100 (46) | 2.95 | 1.52 - 5.7 | 0.0013 |
| COMPERA2 > 2.2 | 82 (32) | 3.5 | 1.5 - 8.17 | 0.0037 |
| REVEAL Lite 2 > 7 | 75 (32) | 1.76 | 0.82 - 3.76 | 0.1449 |
| COMPERA2_Code > 2 | 82 (32) | 3.33 | 1.49 - 7.43 | 0.0033 |
| REVEAL Lite 2 Code > 1 | 75 (32) | 1.76 | 0.82 - 3.76 | 0.1449 |
| COMPERA2 + PHIHDL > 0.5 | 82 (32) | 3.39 | 1.49 - 7.74 | 0.0037 |
| REVEAL Lite 2 + PHIHDL > 0.5 | 75 (32) | 2.41 | 1.12 - 5.19 | 0.0252 |
| COMPERA2 Code + PHIHDL > 0.5 | 82 (32) | 3.36 | 1.47 - 7.67 | 0.004 |
| REVEAL Lite 2 Code + PHIHDL > 0.6 | 75 (32) | 2.21 | 1.02 - 4.81 | 0.0453 |
| COMPERA2 BswP > 2.2 | 82 (32) | 3.02 | 1.39 - 6.55 | 0.0052 |
| REVEAL Lite 2 BswP > 6.5 | 75 (32) | 2.39 | 1.13 - 5.06 | 0.0225 |
| COMPERA2 Code BswP > 2 | 82 (32) | 3.02 | 1.39 - 6.55 | 0.0052 |
| REVEAL Lite 2 Code BswP > 1 | 75 (32) | 2.39 | 1.13 - 5.06 | 0.0225 |

**Table S5:**
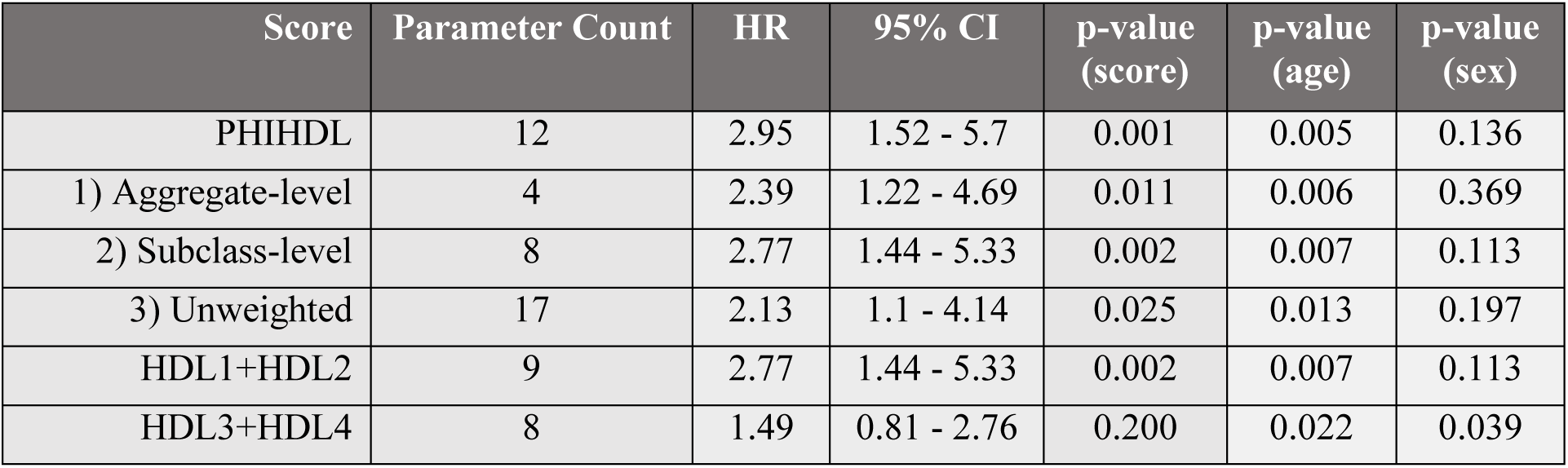
Sensitivity analysis of PHIHDL. Different Cox models were tested to investigate the impact of overlapping/redundant parameters in PHIHDL, and to investigate the role of large and small HDL.

| Score | Parameter Count | HR | 95% CI | p-value<br>(score) | p-value<br>(age) | p-value<br>(sex) |
| --- | --- | --- | --- | --- | --- | --- |
| PHIHDL | 12 | 2.95 | 1.52 - 5.7 | 0.001 | 0.005 | 0.136 |
| 1) Aggregate-level | 4 | 2.39 | 1.22 - 4.69 | 0.011 | 0.006 | 0.369 |
| 2) Subclass-level | 8 | 2.77 | 1.44 - 5.33 | 0.002 | 0.007 | 0.113 |
| 3) Unweighted | 17 | 2.13 | 1.1 - 4.14 | 0.025 | 0.013 | 0.197 |
| HDL1+HDL2 | 9 | 2.77 | 1.44 - 5.33 | 0.002 | 0.007 | 0.113 |
| HDL3+HDL4 | 8 | 1.49 | 0.81 - 2.76 | 0.200 | 0.022 | 0.039 |

## Supplementary Figures

**Fig. S1.**
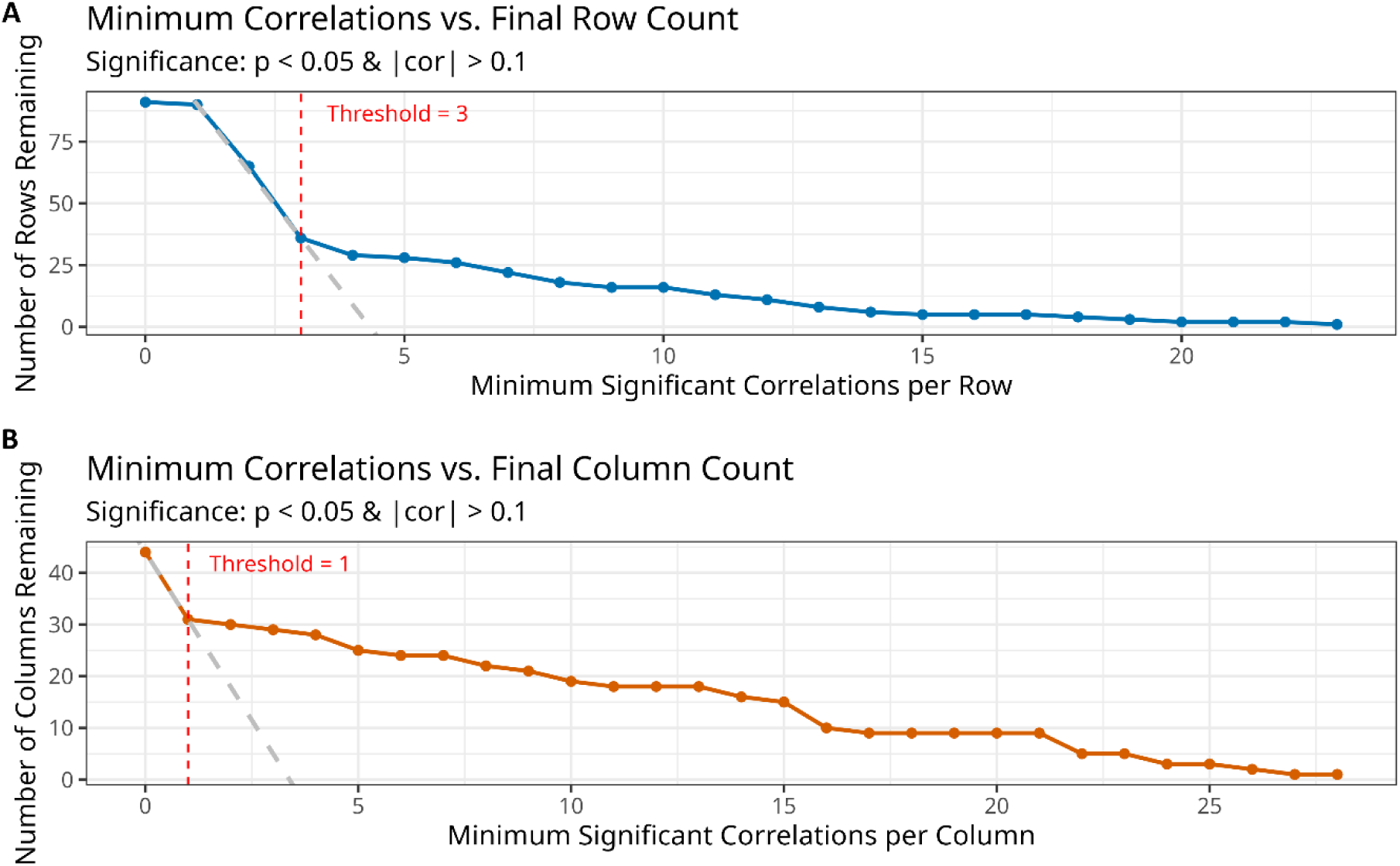
Threshold determination for heatmap filtering. **(A)** Count of remaining metabolic parameters (y-axis) as a function of the minimum number of significant correlations required for each parameter to be kept (x-axis). **(B)** Count of remaining clinical parameters (y-axis) as a function of the minimum number of significant correlations required for each parameter to be kept (x-axis). The dashed lines extrapolate the initial linear relationship. The final threshold is selected at the point where the data curve (blue/orange) diverges from this extrapolated line (grey). In Figure A, x=1 was used as the starting point since almost all metabolic parameters correlated with at least one clinical parameter.

**Fig. S2.**
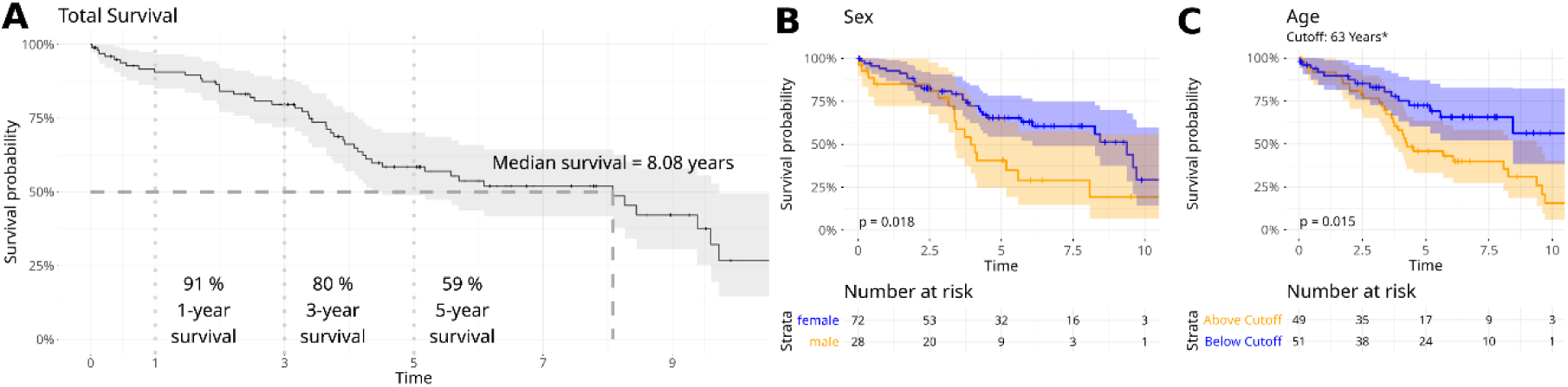
Univariate cohort overview. **(A)** Kaplan-Meier curve showcasing 1-, 3-, and 5-year survival as well as median survival. **(B)** Kaplan-Meier curve stratified by sex. **(C)** Kaplan-Meier curve stratified by age using the cohort median.

**Fig. S3.**
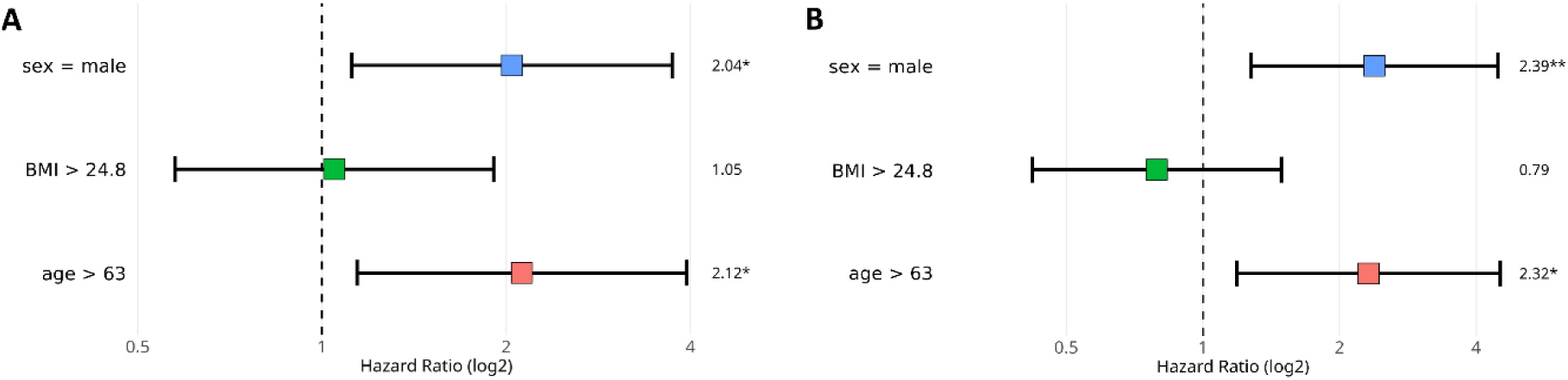
Hazard ratios of sex, BMI and age as potential covariates for correction. **A)** Univariable Cox proportional hazards models for sex, BMI, and age, fitted separately. **B)** Multivariable Cox proportional hazards model including sex, BMI, and age simultaneously. Hazard ratios are shown with 95% confidence intervals.

**Fig. S4.**
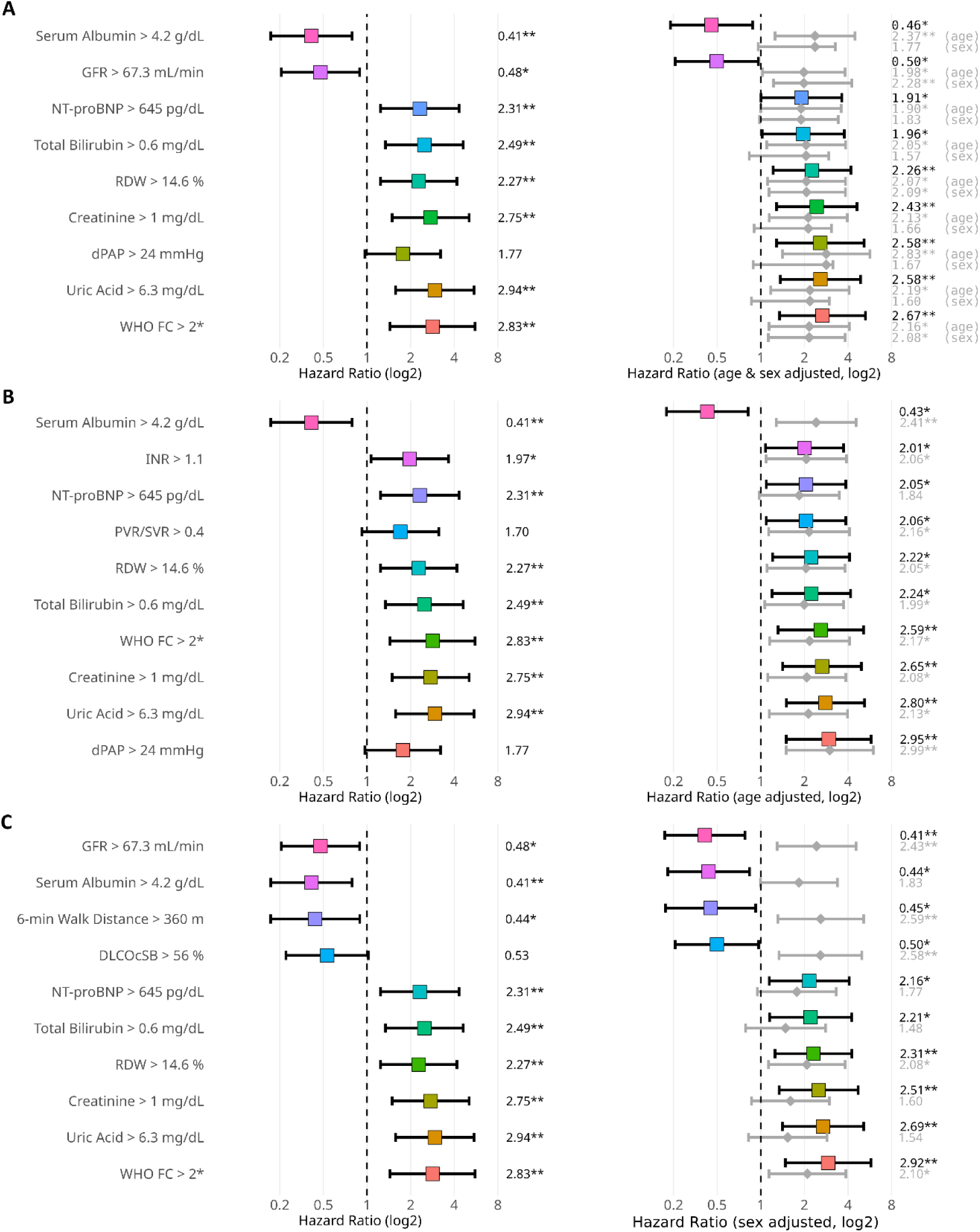
Significant hazard ratios of clinical parameters. **(A)** Hazard ratio adjusted for both age and sex. **(B)** Hazard ratio adjusted for age only. **(C)** Hazard ratio adjusted for sex only. Colored bars indicate confidence intervals for the scores themselves, whereas the grey bars beneath concern the respective confounder (age, sex). * Asterisks indicate that the threshold for dichotomization was set manually rather than using the median to guarantee equal group sizes.

**Fig. S5.**
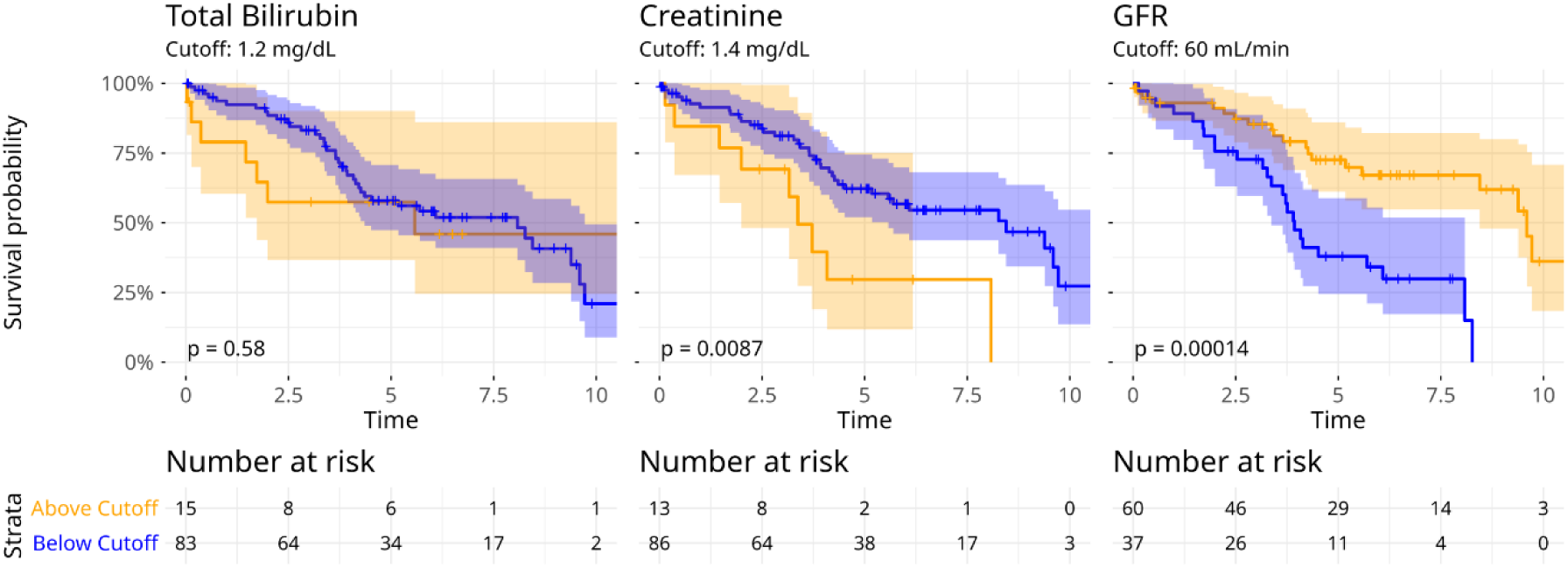
Univariate Kaplan-Meier curves stratified by general survival-related biomarkers using thresholds found in literature. (**25,55,56**)

**Fig. S6:**
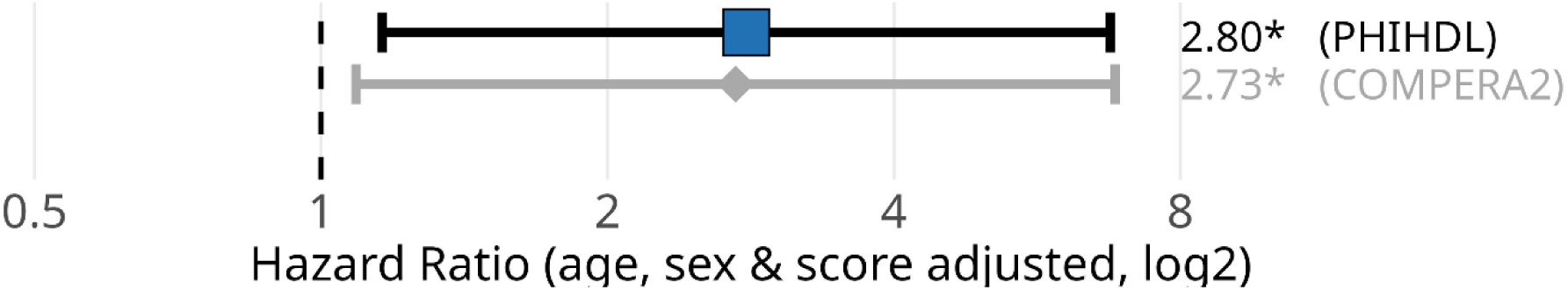
Cox-Proportional-Hazard models when correcting PHIHDL and COMPERA2 for one another. Additive Cox-Proportional hazard for PHIHDL and COMPERA2, additionally adjusted for age and sex (not explicitly shown).

**Fig. S7.**
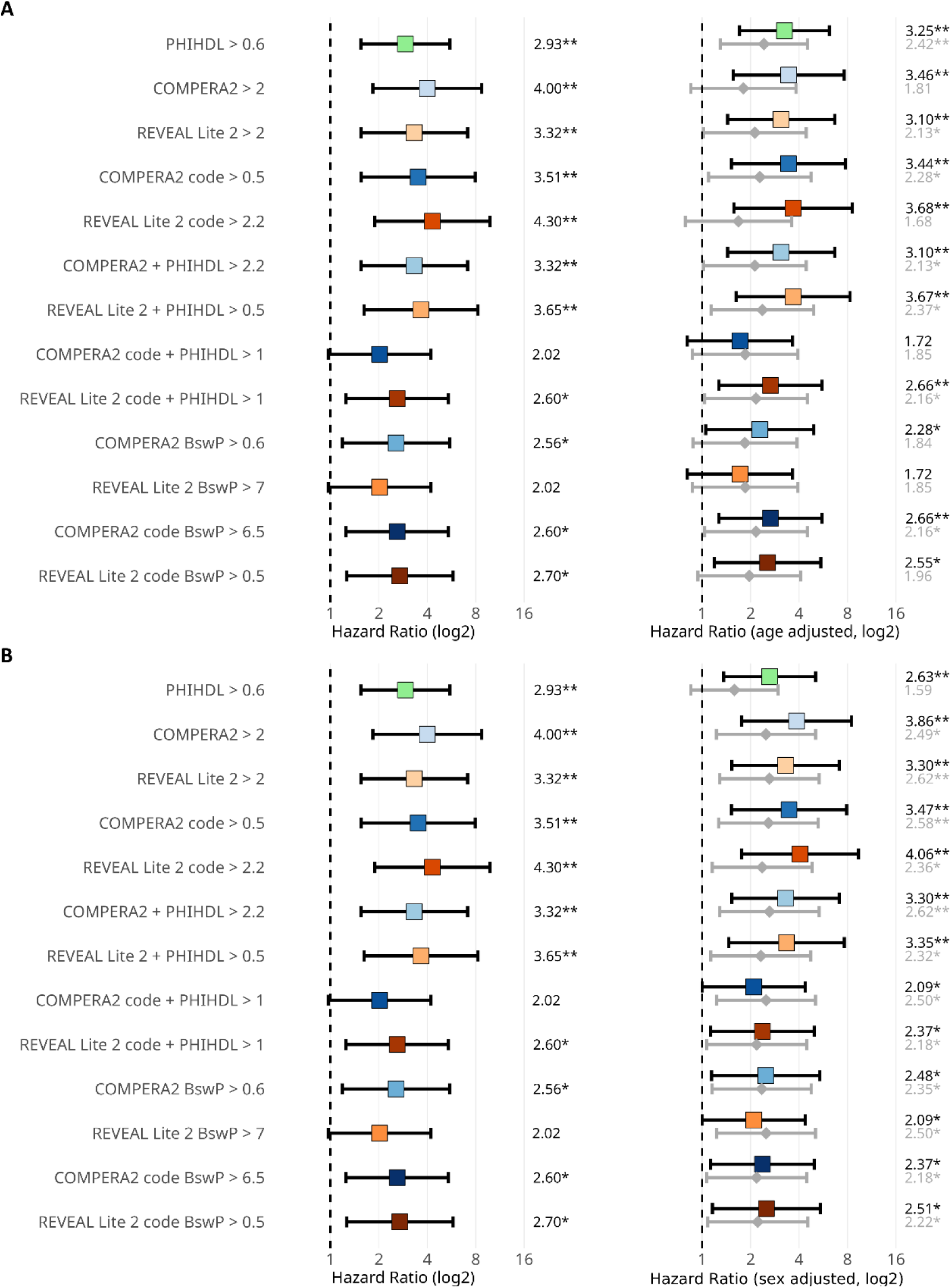
Scores adjusted for one covariate only. **(A)** Scores adjusted for age only. **(B)** Scores adjusted for sex only. Colored bars indicate confidence intervals for the scores themselves, whereas the grey bars represent the respective confounder (in A age, in B sex).

**Fig. S8.**
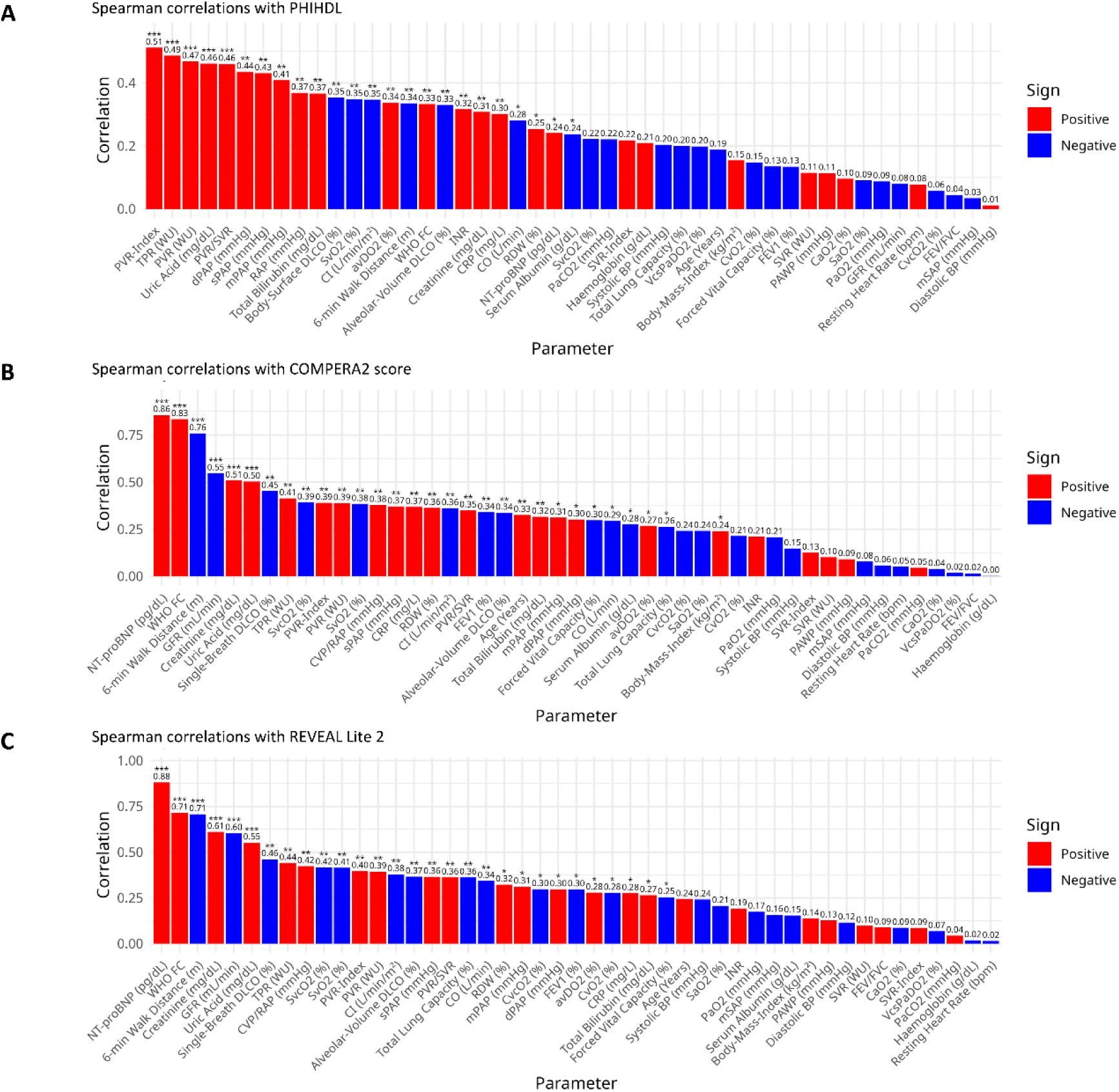
Decomposition of prognostic scores based on their Spearman correlation with various clinical and hemodynamic parameters. **(A)** Correlations with PHIHDL. **(B) C**orrelations with COMPERA2 score and **(C)** correlations with REVEAL Lite 2.

**Fig. S9.**
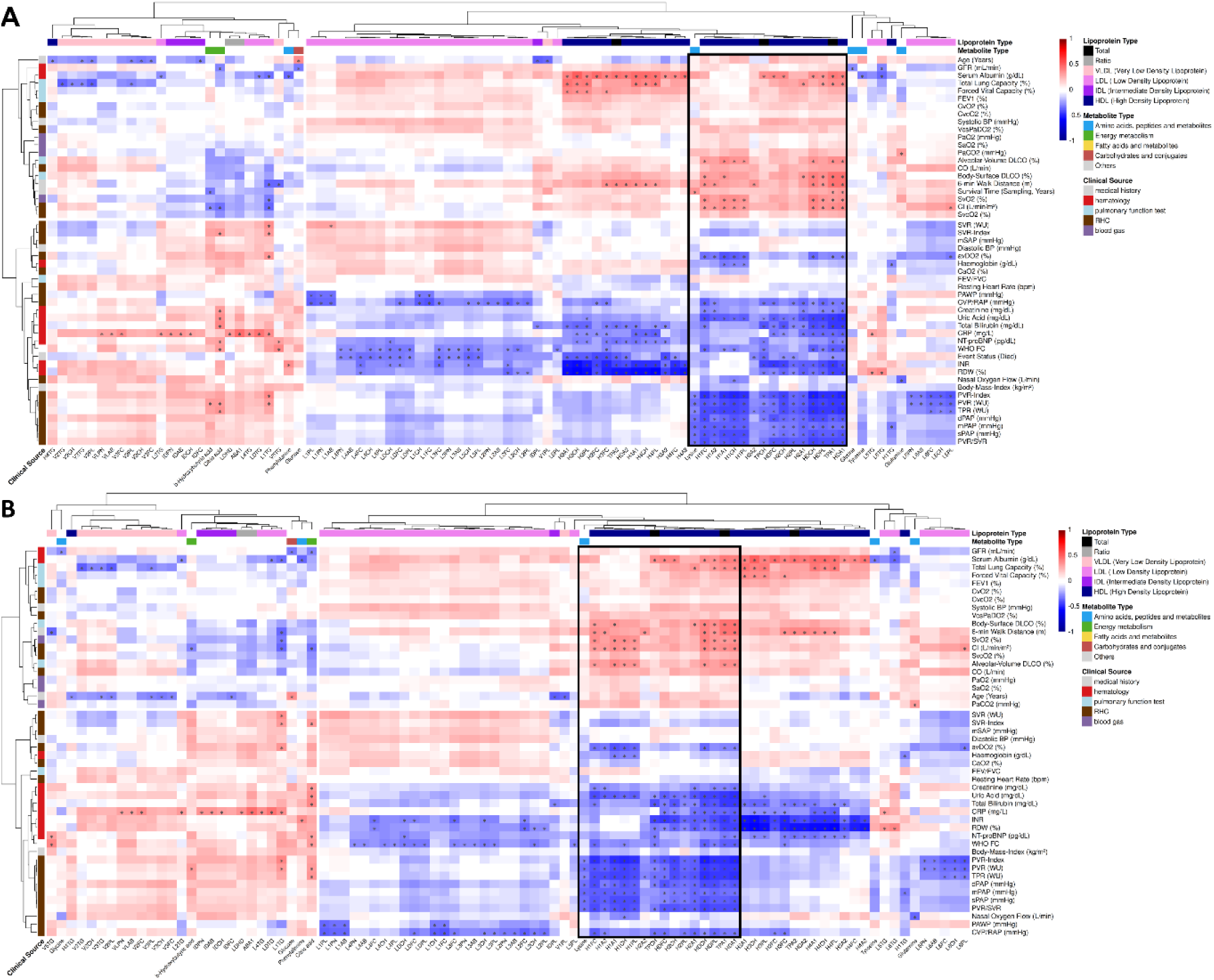
Correlation heatmap for the identification of PHIHDL. **(A)** Original heatmap showcasing correlation between all metabolic parameters containing at least one correlation with clinical values. **(B)** Sensitivity analysis shows that the HDL cluster remains unchanged even after excluding survival data.

**Fig. S10.**
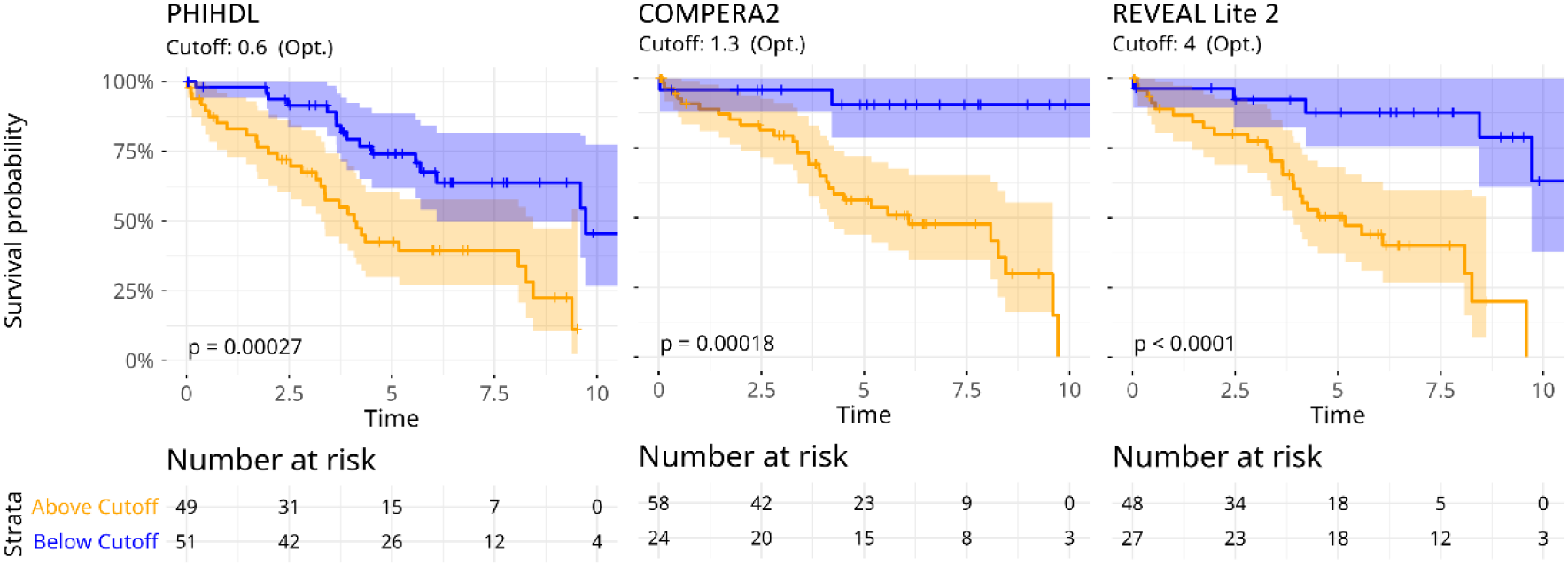
Univariate Kaplan-Meier survival analysis of PAH patients stratified by PHIHDL, COMPERA 2.0, and REVEAL Lite 2 scores using optimized binary stratification (R surv_cutpoint function; minimum 30% group size).

**Fig. S11.**
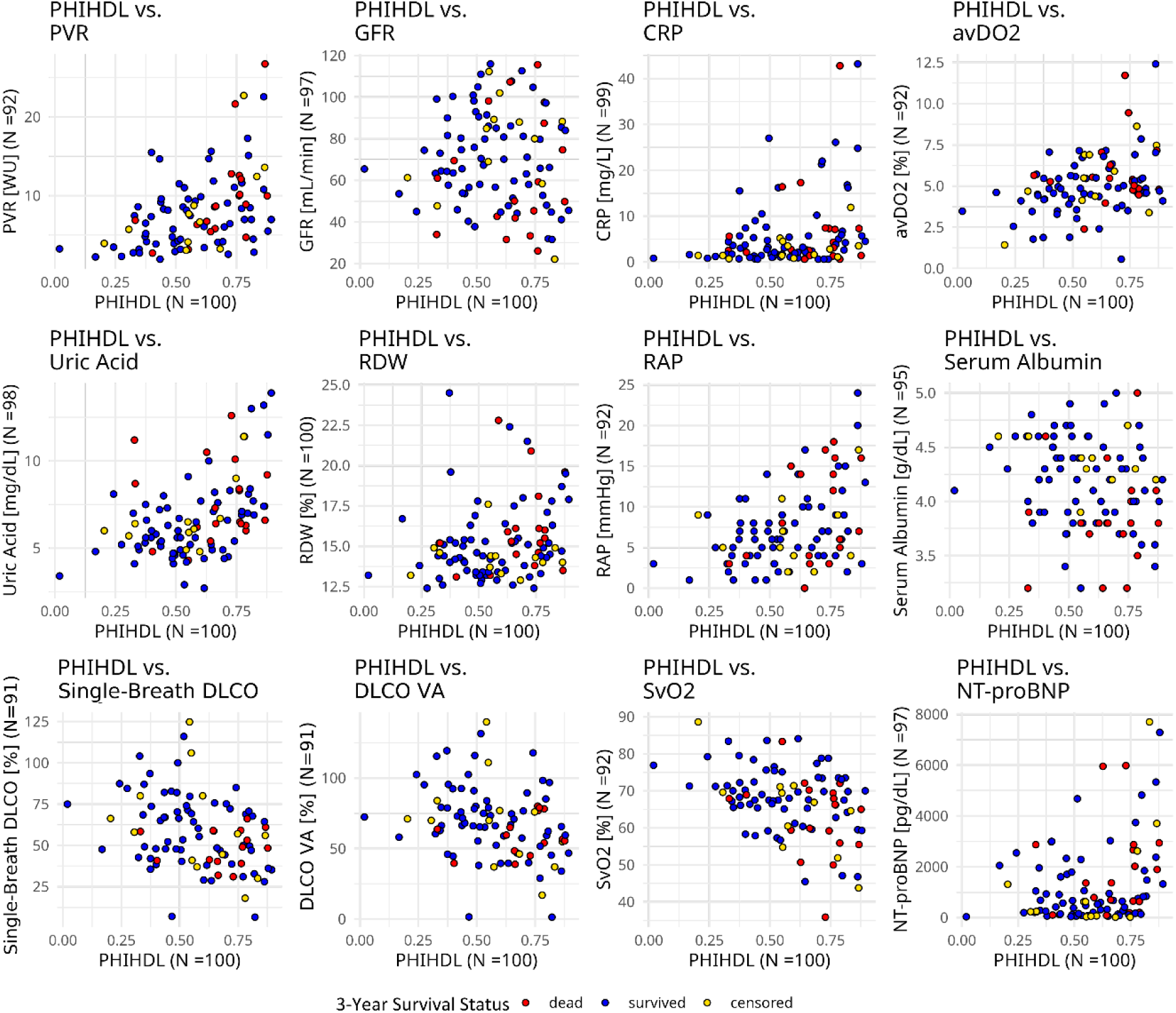
Scatter plot of 3-year survival for combinations of PHIHDL and selected clinical parameters, also showing the amount of samples with populated values (pairwise comparison). In the plots of PHIHDL vs. DLCO, most of the red dots are in the lower right quadrant. This suggests that the combination of high PHIHDL (i.e. low HDL) with low DLCO is most predictive of poor 3-yr survival. The same applies for the combination of high PHIHDL and low SvO2 and low albumin, respectively. DLCO VA, Single-Breath DLCO per alveolar volume, % predicted.

## References

1. Kovacs G, Bartolome S, Denton CP, Gatzoulis MA, Gu S, Khanna D, et al. Definition, classification and diagnosis of pulmonary hypertension. Eur Respir J. 2024 Oct;64(4):2401324. doi:10.1183/13993003.01324-2024 PubMed PMID: 39209475; PubMed Central PMCID: PMC11533989.

2. Chin KM, Gaine SP, Gerges C, Jing ZC, Mathai SC, Tamura Y, et al. Treatment algorithm for pulmonary arterial hypertension. Eur Respir J. 2024 Oct;64(4):2401325. doi:10.1183/13993003.01325-2024 PubMed PMID: 39209476; PubMed Central PMCID: PMC11525349.

3. Guignabert C, Aman J, Bonnet S, Dorfmüller P, Olschewski AJ, Pullamsetti S, et al. Pathology and pathobiology of pulmonary hypertension: current insights and future directions. Eur Respir J. 2024 Oct;64(4):2401095. doi:10.1183/13993003.01095-2024 PubMed PMID: 39209474; PubMed Central PMCID: PMC11533988.

4. Asosingh K, Aldred MA, Vasanji A, Drazba J, Sharp J, Farver C, et al. Circulating angiogenic precursors in idiopathic pulmonary arterial hypertension. Am J Pathol. 2008 Mar;172(3):615–27. doi:10.2353/ajpath.2008.070705 PubMed PMID: 18258847; PubMed Central PMCID: PMC2258264.

5. Toshner M, Voswinckel R, Southwood M, Al-Lamki R, Howard LSG, Marchesan D, et al. Evidence of Dysfunction of Endothelial Progenitors in Pulmonary Arterial Hypertension. Am J Respir Crit Care Med. 2009 Oct 15;180(8):780–7. doi:10.1164/rccm.200810-1662OC PubMed PMID: 19628780; PubMed Central PMCID: PMC2778151.

6. Yeager ME, Nguyen CM, Belchenko DD, Colvin KL, Takatsuki S, Ivy DD, et al. Circulating myeloid-derived suppressor cells are increased and activated in pulmonary hypertension. Chest. 2012 Apr;141(4):944–52. doi:10.1378/chest.11-0205 PubMed PMID: 21940769; PubMed Central PMCID: PMC3318947.

7. Foris V, Kovacs G, Marsh LM, Bálint Z, Tötsch M, Avian A, et al. CD133+ cells in pulmonary arterial hypertension. Eur Respir J. 2016 Aug;48(2):459–69. doi:10.1183/13993003.01523-2015 PubMed PMID: 27103380.

8. Rhodes CJ, Howard LS, Busbridge M, Ashby D, Kondili E, Gibbs JSR, et al. Iron deficiency and raised hepcidin in idiopathic pulmonary arterial hypertension: clinical prevalence, outcomes, and mechanistic insights. J Am Coll Cardiol. 2011 Jul 12;58(3):300–9. doi:10.1016/j.jacc.2011.02.057 PubMed PMID: 21737024.

9. Savale L, Akagi S, Tu L, Cumont A, Thuillet R, Phan C, et al. Serum and pulmonary uric acid in pulmonary arterial hypertension. Eur Respir J. 2021 Aug;58(2):2000332. doi:10.1183/13993003.00332-2020 PubMed PMID: 33446602.

10. Hemnes AR, Luther JM, Rhodes CJ, Burgess JP, Carlson J, Fan R, et al. Human PAH is characterized by a pattern of lipid-related insulin resistance. JCI Insight. 2019 Jan 10;4(1):e123611. doi:10.1172/jci.insight.123611 PubMed PMID: 30626738; PubMed Central PMCID: PMC6485674.

11. Nagy BM, Kovacs G, Tornyos A, Svehlikova E, Foris V, Nagaraj C, et al. No indication of insulin resistance in idiopathic pulmonary arterial hypertension with preserved physical activity. Eur Respir J. 2020 Jun;55(6):1901228. doi:10.1183/13993003.01228-2019 PubMed PMID: 32217652.

12. Mey JT, Hari A, Axelrod CL, Fealy CE, Erickson ML, Kirwan JP, et al. Lipids and ketones dominate metabolism at the expense of glucose control in pulmonary arterial hypertension: a hyperglycaemic clamp and metabolomics study. Eur Respir J. 2020 Apr;55(4):1901700. doi:10.1183/13993003.01700-2019 PubMed PMID: 32108049; PubMed Central PMCID: PMC7263739.

13. Bordag N, Nagy BM, Zügner E, Ludwig H, Foris V, Nagaraj C, et al. Lipid Ratios for Diagnosis and Prognosis of Pulmonary Hypertension. Am J Respir Crit Care Med. 2025 Jul;211(7):1264–76. doi:10.1164/rccm.202407-1345OC

14. Zhao QH, Peng FH, Wei H, He J, Chen FD, Di RM, et al. Serum high-density lipoprotein cholesterol levels as a prognostic indicator in patients with idiopathic pulmonary arterial hypertension. Am J Cardiol. 2012 Aug 1;110(3):433–9. doi:10.1016/j.amjcard.2012.03.042 PubMed PMID: 22560769.

15. Heresi GA, Aytekin M, Newman J, DiDonato J, Dweik RA. Plasma Levels of High-Density Lipoprotein Cholesterol and Outcomes in Pulmonary Arterial Hypertension. Am J Respir Crit Care Med. 2010 Sep 1;182(5):661–8. doi:10.1164/rccm.201001-0007OC PubMed PMID: 20448092; PubMed Central PMCID: PMC2937236.

16. Harbaum L, Ghataorhe P, Wharton J, Jiménez B, Howard LSG, Gibbs JSR, et al. Reduced plasma levels of small HDL particles transporting fibrinolytic proteins in pulmonary arterial hypertension. Thorax. 2019 Apr 1;74(4):380–9. doi:10.1136/thoraxjnl-2018-212144 PubMed PMID: 30478197.

17. Humbert M, Kovacs G, Hoeper MM, Badagliacca R, Berger RMF, Brida M, et al. 2022 ESC/ERS Guidelines for the diagnosis and treatment of pulmonary hypertension. Eur Heart J. 2022 Oct 11;43(38):3618–731. doi:10.1093/eurheartj/ehac237 PubMed PMID: 36017548.

18. Stadler JT, Borenich A, Pammer A, Emrich IE, Habisch H, Madl T, et al. Association of Small HDL Subclasses with Mortality Risk in Chronic Kidney Disease. Antioxid Basel Switz. 2024 Dec 11;13(12):1511. doi:10.3390/antiox13121511 PubMed PMID: 39765838; PubMed Central PMCID: PMC11673888.

19. Abdi H, Williams LJ. Principal component analysis. WIREs Comput Stat. 2010;2(4):433–59. doi:10.1002/wics.101

20. Benjamini Y, Hochberg Y. Controlling the False Discovery Rate: A Practical and Powerful Approach to Multiple Testing. J R Stat Soc Ser B Methodol. 1995;57(1):289–300. doi:10.1111/j.2517-6161.1995.tb02031.x

21. Cattell RB. The Scree Test For The Number Of Factors. Multivar Behav Res. 1966 Apr 1;1(2):245– 76. doi:10.1207/s15327906mbr0102_10 PubMed PMID: 26828106.

22. Lewis F, Butler A, Gilbert L. A unified approach to model selection using the likelihood ratio test. Methods Ecol Evol. 2011;2(2):155–62. doi:10.1111/j.2041-210X.2010.00063.x

23. Akaike H. Akaike’s Information Criterion. In: International Encyclopedia of Statistical Science [Internet]. Springer, Berlin, Heidelberg; 2025 [cited 2026 May 12]. p. 41–2. Available from: https://link.springer.com/rwe/10.1007/978-3-662-69359-9_14 doi:10.1007/978-3-662-69359-9_14

24. GRAMBSCH PM, THERNEAU TM. Proportional hazards tests and diagnostics based on weighted residuals. Biometrika. 1994 Sep 1;81(3):515–26. doi:10.1093/biomet/81.3.515

25. Dardi F, Boucly A, Benza R, Frantz R, Mercurio V, Olschewski H, et al. Risk stratification and treatment goals in pulmonary arterial hypertension. Eur Respir J. 2024 Oct;64(4):2401323. doi:10.1183/13993003.01323-2024 PubMed PMID: 39209472; PubMed Central PMCID: PMC11525341.

26. Larsen CM, McCully RB, Murphy JG, Kushwaha SS, Frantz RP, Kane GC. Usefulness of High-Density Lipoprotein Cholesterol to Predict Survival in Pulmonary Arterial Hypertension. Am J Cardiol. 2016 Jul 15;118(2):292–7. doi:10.1016/j.amjcard.2016.04.028 PubMed PMID: 27291969.

27. Wang GF, Guan LH, Zhou DX, Chen DD, Zhang XC, Ge JB. Serum High-Density Lipoprotein Cholesterol is Significantly Associated with the Presence and Severity of Pulmonary Arterial Hypertension: A Retrospective Cross-Sectional Study. Adv Ther. 2020 May;37(5):2199–209. doi:10.1007/s12325-020-01304-2 PubMed PMID: 32239458.

28. Cracowski JL, Labarère J, Renversez JC, Degano B, Chabot F, Humbert M. Plasma levels of high-density lipoprotein cholesterol are not associated with survival in pulmonary arterial hypertension. Am J Respir Crit Care Med. 2012 Jul 1;186(1):107; author reply 107-108. doi:10.1164/ajrccm.186.1.107 PubMed PMID: 22753690.

29. Hoeper MM, Pausch C, Olsson KM, Huscher D, Pittrow D, Grünig E, et al. COMPERA 2.0: a refined four-stratum risk assessment model for pulmonary arterial hypertension. Eur Respir J. 2022 Jul 7;60(1). doi:10.1183/13993003.02311-2021 PubMed PMID: 34737226.

30. Hansmann G, de Jesus Perez VA, Alastalo TP, Alvira CM, Guignabert C, Bekker JM, et al. An antiproliferative BMP-2/PPARã/apoE axis in human and murine SMCs and its role in pulmonary hypertension. J Clin Invest. 2008 May 1;118(5):1846–57. doi:10.1172/JCI32503 PubMed PMID: 18382765; PubMed Central PMCID: PMC2276393.

31. Hansmann G, Wagner RA, Schellong S, Perez VA de J, Urashima T, Wang L, et al. Pulmonary arterial hypertension is linked to insulin resistance and reversed by peroxisome proliferator-activated receptor-gamma activation. Circulation. 2007 Mar 13;115(10):1275–84. doi:10.1161/CIRCULATIONAHA.106.663120 PubMed PMID: 17339547.

32. Lawrie A, Hameed AG, Chamberlain J, Arnold N, Kennerley A, Hopkinson K, et al. Paigen diet-fed apolipoprotein E knockout mice develop severe pulmonary hypertension in an interleukin-1-dependent manner. Am J Pathol. 2011 Oct;179(4):1693–705. doi:10.1016/j.ajpath.2011.06.037 PubMed PMID: 21835155; PubMed Central PMCID: PMC3181351.

33. Brittain EL, Talati M, Fessel JP, Zhu H, Penner N, Calcutt MW, et al. Fatty Acid Metabolic Defects and Right Ventricular Lipotoxicity in Human Pulmonary Arterial Hypertension. Circulation. 2016 May 17;133(20):1936–44. doi:10.1161/CIRCULATIONAHA.115.019351

34. Lewis GD, Ngo D, Hemnes AR, Farrell L, Domos C, Pappagianopoulos PP, et al. Metabolic Profiling of Right Ventricular-Pulmonary Vascular Function Reveals Circulating Biomarkers of Pulmonary Hypertension. J Am Coll Cardiol. 2016 Jan 19;67(2):174–89. doi:10.1016/j.jacc.2015.10.072 PubMed PMID: 26791065; PubMed Central PMCID: PMC4962613.

35. Nagy BM, Nagaraj C, Meinitzer A, Sharma N, Papp R, Foris V, et al. Importance of kynurenine in pulmonary hypertension. Am J Physiol Lung Cell Mol Physiol. 2017 Nov 1;313(5):L741–51. doi:10.1152/ajplung.00517.2016 PubMed PMID: 28705908.

36. Gordon DJ, Probstfield JL, Garrison RJ, Neaton JD, Castelli WP, Knoke JD, et al. High-density lipoprotein cholesterol and cardiovascular disease. Four prospective American studies. Circulation. 1989 Jan;79(1):8–15. doi:10.1161/01.CIR.79.1.8

37. Tall AR, Yvan-Charvet L, Terasaka N, Pagler T, Wang N. HDL, ABC transporters, and cholesterol efflux: implications for the treatment of atherosclerosis. Cell Metab. 2008 May;7(5):365–75. doi:10.1016/j.cmet.2008.03.001 PubMed PMID: 18460328.

38. Nofer JR, Assmann G. Atheroprotective Effects of High-Density Lipoprotein-Associated Lysosphingolipids. Trends Cardiovasc Med. 2005 Oct 1;15(7):265–71. doi:10.1016/j.tcm.2005.08.005

39. Tölle M, Pawlak A, Schuchardt M, Kawamura A, Tietge UJ, Lorkowski S, et al. HDL-associated lysosphingolipids inhibit NAD(P)H oxidase-dependent monocyte chemoattractant protein-1 production. Arterioscler Thromb Vasc Biol. 2008 Aug;28(8):1542–8. doi:10.1161/ATVBAHA.107.161042 PubMed PMID: 18483405; PubMed Central PMCID: PMC2723752.

40. Li XP, Zhao SP, Zhang XY, Liu L, Gao M, Zhou QC. Protective effect of high density lipoprotein on endothelium-dependent vasodilatation. Int J Cardiol. 2000 May 31;73(3):231–6. doi:10.1016/s0167-5273(00)00221-7 PubMed PMID: 10841964.

41. Kuvin JT, Patel AR, Sidhu M, Rand WM, Sliney KA, Pandian NG, et al. Relation between high-density lipoprotein cholesterol and peripheral vasomotor function. Am J Cardiol. 2003 Aug 1;92(3):275–9. doi:10.1016/s0002-9149(03)00623-4 PubMed PMID: 12888130.

42. Zeiher AM, Schächlinger V, Hohnloser SH, Saurbier B, Just H. Coronary atherosclerotic wall thickening and vascular reactivity in humans. Elevated high-density lipoprotein levels ameliorate abnormal vasoconstriction in early atherosclerosis. Circulation. 1994 Jun;89(6):2525–32. doi:10.1161/01.CIR.89.6.2525

43. Bisoendial RJ, Hovingh GK, Levels JHM, Lerch PG, Andresen I, Hayden MR, et al. Restoration of Endothelial Function by Increasing High-Density Lipoprotein in Subjects With Isolated Low High-Density Lipoprotein. Circulation. 2003 Jun 17;107(23):2944–8. doi:10.1161/01.CIR.0000070934.69310.1A

44. Podrez EA. Anti-oxidant properties of high-density lipoprotein and atherosclerosis. Clin Exp Pharmacol Physiol. 2010 Jul;37(7):719–25. doi:10.1111/j.1440-1681.2010.05380.x PubMed PMID: 20374263; PubMed Central PMCID: PMC3010184.

45. Mahley RW, Huang Y, Weisgraber KH. Putting cholesterol in its place: apoE and reverse cholesterol transport. J Clin Invest. 2006 May 1;116(5):1226–9. doi:10.1172/JCI28632 PubMed PMID: 16670767.

46. Rader DJ, Hovingh GK. HDL and cardiovascular disease. Lancet Lond Engl. 2014 Aug 16;384(9943):618–25. doi:10.1016/S0140-6736(14)61217-4 PubMed PMID: 25131981.

47. Rohatgi A, Khera A, Berry JD, Givens EG, Ayers CR, Wedin KE, et al. HDL cholesterol efflux capacity and incident cardiovascular events. N Engl J Med. 2014 Dec 18;371(25):2383–93. doi:10.1056/NEJMoa1409065 PubMed PMID: 25404125; PubMed Central PMCID: PMC4308988.

48. Wishart DS, Guo A, Oler E, Wang F, Anjum A, Peters H, et al. HMDB 5.0: the Human Metabolome Database for 2022. Nucleic Acids Res. 2022 Jan 7;50(D1):D622–31. doi:10.1093/nar/gkab1062 PubMed PMID: 34986597; PubMed Central PMCID: PMC8728138.

49. Leys C, Ley C, Klein O, Bernard P, Licata L. Detecting outliers: Do not use standard deviation around the mean, use absolute deviation around the median. J Exp Soc Psychol. 2013 Jul 1;49(4):764–6. doi:10.1016/j.jesp.2013.03.013

50. Emwas AH, Roy R, McKay RT, Tenori L, Saccenti E, Gowda GAN, et al. NMR Spectroscopy for Metabolomics Research. Metabolites. 2019 Jun 27;9(7):123. doi:10.3390/metabo9070123 PubMed PMID: 31252628; PubMed Central PMCID: PMC6680826.

51. Broadhurst DI, Kell DB. Statistical strategies for avoiding false discoveries in metabolomics and related experiments. Metabolomics. 2006 Dec 1;2(4):171–96. doi:10.1007/s11306-006-0037-z

52. Cohen J. Statistical Power Analysis for the Behavioral Sciences. 2nd edn. New York: Routledge; 2013. 567 p. doi:10.4324/9780203771587

53. Mazer NA, Giulianini F, Paynter NP, Jordan P, Mora S. A comparison of the theoretical relationship between HDL size and the ratio of HDL cholesterol to apolipoprotein A-I with experimental results from the Women’s Health Study. Clin Chem. 2013 Jun;59(6):949–58. doi:10.1373/clinchem.2012.196949 PubMed PMID: 23426429; PubMed Central PMCID: PMC3669243.

54. Walldius G, Jungner I. The apoB/apoA-I ratio: a strong, new risk factor for cardiovascular disease and a target for lipid-lowering therapy--a review of the evidence. J Intern Med. 2006 May;259(5):493–519. doi:10.1111/j.1365-2796.2006.01643.x PubMed PMID: 16629855.

55. Benza RL, Gomberg-Maitland M, Elliott CG, Farber HW, Foreman AJ, Frost AE, et al. Predicting Survival in Patients With Pulmonary Arterial Hypertension: The REVEAL Risk Score Calculator 2.0 and Comparison With ESC/ERS-Based Risk Assessment Strategies. Chest. 2019 Aug 1;156(2):323–37. doi:10.1016/j.chest.2019.02.004

56. Takeda Y, Takeda Y, Tomimoto S, Tani T, Narita H, Kimura G. Bilirubin as a prognostic marker in patients with pulmonary arterial hypertension. BMC Pulm Med. 2010 Apr 22;10:22. doi:10.1186/1471-2466-10-22 PubMed PMID: 20412580; PubMed Central PMCID: PMC2873470.

